# Efficacy of ultra-short, response-guided sofosbuvir and daclatasvir therapy for Hepatitis C: a single arm mechanistic pilot study

**DOI:** 10.1101/2022.08.15.22278752

**Authors:** Barnaby Flower, Le Manh Hung, Leanne McCabe, M. Azim Ansari, Chau Le Ngoc, Thu Vo Thi, Hang Vu Thi Kim, Phuong Nguyen Thi Ngoc, Le Thanh Phuong, Vo Minh Quang, Thuan Dang Trong, Thao Le Thi, Tran Nguyen Bao, Cherry Kingsley, David Smith, Richard M. Hoglund, Joel Tarning, Evelyne Kestelyn, Sarah L Pett, Rogier van Doorn, Jennifer Ilo van Nuil, Hugo Turner, Guy Thwaites, Eleanor Barnes, Motiur Rahman, Ann Sarah Walker, Jeremy Day, Nguyen Van Vinh Chau, Graham S Cooke

## Abstract

**Background:** WHO has called for research into predictive factors for selecting persons who could be successfully treated with shorter durations of direct acting antiviral (DAA) therapy for Hepatitis C. We evaluated early virological response as a means of shortening treatment and explored host, viral and pharmacokinetic contributors to treatment outcome.

**Methods:** Duration of sofosbuvir and daclatasvir (SOF/DCV) was determined according to day 2 (D2) virologic response for HCV genotype (gt) 1- or 6-infected adults in Vietnam with mild liver disease. Participants received 4 or 8 weeks treatment according to whether D2 HCV RNA was above or below 500 IU/ml (standard duration is 12 weeks). Primary endpoint was sustained virological response (SVR12). Those failing therapy were retreated with 12 weeks SOF/DCV. Host IFNL4 genotype and viral sequencing was performed at baseline, with repeat viral sequencing if virological rebound was observed. Levels of SOF, its inactive metabolite GS-331007 and DCV were measured on day 0 and 28.

**Results:** Of 52 adults enrolled, 34 received 4 weeks SOF/DCV, 17 got 8 weeks and one withdrew. SVR12 was achieved in 21/34 (62%) treated for 4 weeks, and 17/17 (100%) treated for 8 weeks. Overall 38/51 (75%) were cured with first-line treatment (mean duration 37 days). Despite a high prevalence of putative NS5A-inhibitor resistance associated substitutions (RAS), all first-line treatment failures cured after retreatment (13/13). We found no evidence treatment failure was associated with host IFNL4 genotype, viral subtype, baseline RAS or DCV levels. SOF metabolite levels were higher in those failing 4-week therapy.

**Conclusions:** Shortened SOF/DCV therapy, with retreatment if needed, reduces DAA use while maintaining high cure rates. D2 virologic response alone does not adequately predict SVR12 with 4 weeks treatment.

**Funding:** Funded by the Medical Research Council (grant MR/P025064/1) and The Global Challenges Research Fund (Wellcome Trust Grant 206/296/Z/17/Z).)

**Clinical trial number:** ISRCTN17100273

## INTRODUCTION

Directly acting antiviral (DAA) therapy for hepatitis C (HCV) offers high cure rates to those able to adhere to standard durations of treatment. In low- and middle-income countries, where treatment is limited to second generation NS5A/NS5B-inhibitor combinations, standard treatment is at least 12 weeks. This duration presents a barrier to successful engagement in care for some populations^1,2^, hampering the elimination of HCV as a public health threat. Novel treatment strategies are required for hard-to-reach individuals such as people who inject drugs and those of no fixed abode.

In Vietnam DAA therapy remains prohibitively expensive for many of those infected. A standard twelve-week course of sofosbuvir and daclatasvir (SOF/DCV) was priced at US$2417 - 2472 in Ho Chi Minh City in 2019^3^. Despite the government subsidising 50% of drug costs since, the Ministry of Health estimate only 1000 individuals accessed DAA treatment through health insurance in 2019, and 2700 in 2020^4^.

The World Health Organisation has called for research into predictive factors for selecting persons who could be successfully treated with shorter durations of therapy^5^, which could expand access to treatment and reduce drug costs. Studies evaluating short course therapy are challenging for infectious diseases where there are significant clinical risks of failure (e.g. TB, Sepsis). However, HCV provides a model where treatment failures can be successfully retreated^6^ allowing exploration of mechanisms underlying successful therapy.

Shortened DAA therapy is generally associated with disappointing rates of cure, such that it could never be recommended routinely. A systematic review and meta-analysis into treatment optimisation for HCV with DAA therapy in individuals with favourable predictors of response, found that pooled sustained virological response (SVR) for regimens of ≤4 weeks duration was 63.1% (95% C.I. 39.9-83.7), 6 weeks duration was 81.1% (75.1-86.6) and 8 weeks duration was 94.2% (92.3-95.9)^7^. However improved rates of cure were seen with an increased number of individual-level factors known (or assumed) to be favourable, such as non-genotype 3 infection, lower body mass index (BMI), lower baseline viral load, mild liver disease, absence of prior treatment failure, and a rapid virological response to treatment^7^.

Rapid virological response offers a promising means of shortening treatment duration while maintaining high rates of cure. So-called response-guided therapy (RGT), whereby antiviral duration is shortened in individuals who rapidly suppress virus levels in blood after starting treatment, was routinely used in the era of interferon-based therapy, when an undetectable HCV RNA at 4 weeks was used to determine a shorter course of pegylated interferon and ribavirin^5^. Evidence supporting RGT with DAAs at earlier timepoints is emerging, notably using day 2 (D2) viral load to determine treatment duration in genotype 1b infection. In this population, high cure rates were observed with just three weeks triple therapy (protease inhibitor, NS5A inhibitor and NS5B inhibitor)^8^. There is currently no data for RGT durations less than 8 weeks with sofosbuvir and daclatasvir (SOF/DCV), which remains the lowest-priced and most widely available treatment option globally^9^.

Drug resistance in association with particular viral genotypes and subtypes is also know to influence treatment outcome^10,11^ and may predict who can be treated with shortened therapy. Vietnam has a high burden of genotype 6 HCV infection (around 35%)^12^, which is rare outside South East Asia and under-represented in clinical trials. Genotype 6 is the most genetically diverse HCV lineage^13^, raising concerns about the potential for emergence of resistant variants^14^.

The human *IFNL4* di-nucleotide polymorphism rs368234815 (ΔG/TT) controls generation of the IFNL4 protein and is also associated with impaired clearance of HCV^15^ and inferior responses to pegylated interferon-alpha/ribavirin therapy^16^ and SOF-based treatment^17,18^. The impact of host *IFNL4* genotype in shortened DAA therapy is not well understood. It is also unknown how serum levels of SOF, its metabolite GS-331007, and DCV might impact treatment success with shortened therapy.

In this prospective single-arm mechanistic study in Ho Chi Minh City (HCMC), individuals with genotypes 1 and 6 HCV infection and mild liver disease were treated with shortened course SOF/DCV. We tested the hypothesis that high rates of cure can be achieved with short course DAAs when early on-treatment virological response is used to guide duration of therapy. We also compared host *IFNL4* genetic polymorphism, DAA drug levels, HCV subtypes and previously defined (in vitro) resistance-associated substitutions in cures versus treatment failures to better understand the biological mechanisms determining treatment outcome.

## METHODS

### Study population

Participants were recruited from the outpatient hepatitis clinic of the Hospital for Tropical Diseases (HTD) in HCMC, between February 2019 and June 2020. Eligible patients were ≥18 years and had chronic infection with HCV genotype 1 or 6 without evidence of liver fibrosis (defined as a FibroScan score ≤7.1kPa, equivalent to F0-F1 disease)^19^. In addition, participants were required to be HCV-treatment naïve, have a BMI ≥18kg/m^2^, a creatinine clearance ≥60ml/min, with no evidence of HIV or Hepatitis B coinfection, or solid organ malignancy in the preceding 5 years. Full eligibility criteria are provided in the protocol available at https://doi.org/10.1186/ISRCTN17100273.

Patients referred to the trial were initially enrolled into an observational study which included fibroscan assessment and genotyping. Individuals in this cohort found to be potentially eligible for the trial were invited for further screening. All patients provided written informed consent.

### Study design

All participants were treated with sofosbuvir 400mg and daclatasvir 60mg (Pharco Pharmaceuticals, Egypt) administered orally as two separate tablets, once daily. Individuals requiring dose adjustment for any reason were excluded.

Treatment duration was determined using hepatitis C viral load measured 2 days after treatment onset (D2). Participants with viral load <500 IU/ml at D2 (after 2 dose of SOF/DCV) were treated with 4 weeks of SOF/DCV. Those with HCV RNA ≥500 IU/ml received 8 weeks (figure 1). This timepoint and viral load threshold were chosen from previous evaluations of response guided therapy with DAAs^8^.

**Figure 1:**
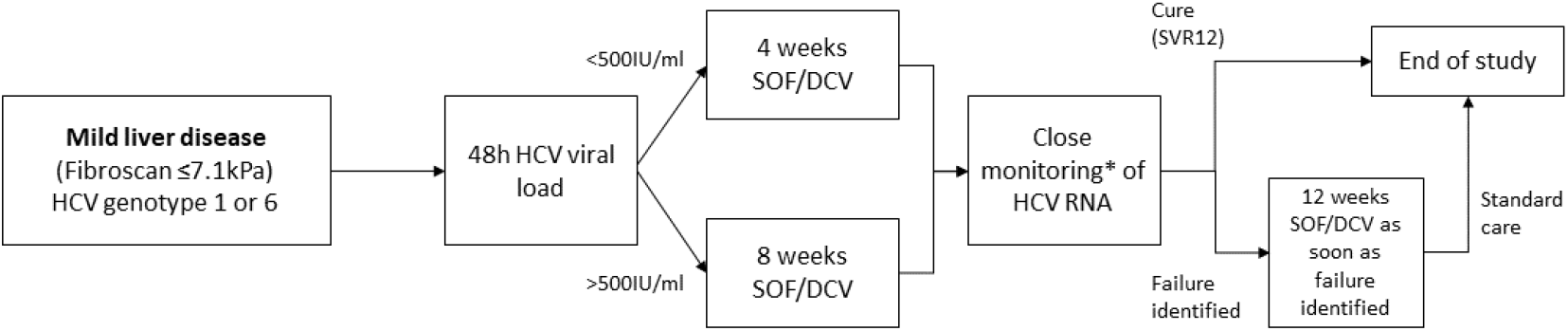
Study design. *HCV RNA on day 0, 1, 2, 7, 10, 14, 17, 21, 24, 28, (42, 56), EOT+3, EOT+7, EOT+10, EOT+14, EOT+17, EOT+21, EOT+24, EOT+28s, EOT+56, EOT+84

To analyse viral kinetics on treatment (and on occasion of any failure), HCV viral load was measured at baseline (day 0) and at all subsequent follow up visits on day 1, 2, 7 and then twice weekly until end of treatment (figure 1). Visits after end-of-treatment (EOT) were scheduled twice weekly in the first month after completion of treatment, and then at 8 and 12 weeks after EOT.

The primary endpoint was sustained virological response (SVR12) defined as plasma HCV RNA less than the lower limit of quantification (<LLOQ) 12 weeks after the end of treatment without prior failure. Failure of first-line treatment was carefully defined to incorporate individuals who fully suppressed HCV RNA (<LLOQ) on therapy with late virological rebound, as well as those who never fully supressed HCV viral load. In both cases two consecutive viral loads >LLOQ, taken at least one week apart, were required to confirm failure, with the second >2000 IU/ml. Once failure was confirmed, participants commenced retreatment with standard duration SOF/DCV within 2 weeks (figure 1).

Secondary endpoints were lack of initial virological response (<1 log10 decrease in HCV viral load from baseline), serious adverse events (SAE), grade 3/4 clinical adverse events (AEs), adverse events of any grade leading to change in treatment (SOF, DCV or any other concomitant medication) and adverse reactions (AR). Severity of all AEs and ARs were graded using the Common Toxicity Criteria for Adverse Events gradings^20^.

### Sample size justification

We set a target cure rate of ≥90%, and an unacceptably low cure rate of 70%. Assuming 90% power and one-sided alpha=0.05, 37 participants were required to exclude the null hypothesis that cure was <90%. Assuming 5% loss to follow-up, and that, based on the study by Lau et al^8^, 65% would suppress viral load <500IU/ml by day 2 and receive 4 weeks (rather than 8 weeks) of therapy, the final target population was 60 participants, pooling genotypes 1 and 6.

### Study assessments

At each visit patients were assessed by a study doctor. AEs and ARs were recorded and graded according to a standardised scale^20^ and medication adherence and use of healthcare facilities were recorded on case report forms.

HCV RNA was measured in the hospital using the available commercial platform. At start of study (for the first 34 participants enrolled), this was the Abbott Architect® (LLOQ = 12 IU/ml). This was subsequently replaced with the COBAS AmpliPrep®/COBAS TaqMan® HCV Quantitative Test, version 2.0 (Roche Molecular Systems, LLOQ = 15 IU/ml). Standard laboratory tests - including full blood count, renal function and liver function tests – were performed in the hospital laboratory at baseline, EOT and EOT+12.

### Virus sequencing

At screening, HCV genotype and subtype were determined using NS5B, Core and 5’ UTR sequencing, according to the method described by Chau et.al^21^. To evaluate the impact of HCV subtypes and resistance associated substitutions on treatment outcome, whole genome sequencing (WGS) was additionally performed on all enrolled participants’ virus at baseline, and upon virological rebound and at start of retreatment in participants failing therapy. WGS of the HCV viral genome was attained using Illumina MiSeq platform as described previously^22–25^. The de novo assemblies nucleotide sequences were translated into amino acid and were aligned to H77 HCV reference (GenBank ID: NC_038882.1) and the NS5A and NS5B protein regions were extracted. We only looked for RAS that were present in at least 15% of the reads in the sample and had a read count of greater than 10.

We used the Public Health England (PHE) HCV Resistance Group’s definition for resistance associated substitutions (RAS)^26^. For genotype 1 we looked for RASs defined specifically for genotype 1 as they are well studied. For genotype 6 we looked for all RASs defined across all genotypes, as little work has been done on RASs in genotype 6.

For DCV we looked for 24R, 28T, 30E/K/T, 31M/V, 32L, 58D, and 93C/H/N/R/S/W in genotype 1 infection and additionally looked for 28S, 30R and 31F in genotype 6 infection. For SOF we looked for 159F, 237G, 282T, 315H/N, 321A/I in genotype 1 infection and additionally looked for 289I in genotype 6 infection^17,18^.

In addition to viral sequencing, we evaluated host genetic polymorphisms within the interferon lambda 4 (*IFNL4*) gene of all participants at baseline. Genotyping of *IFNL4* rs368234815 was performed on host DNA using the TaqMan® SNP genotyping assay and primers described previously^15^ with Type-it Fast SNP Probe PCR Master Mix (Qiagen).

### Pharmacokinetics & pharmacodynamics (PK/PD)

To assess pharmacokinetics (PK) and pharmacodynamics (PD), the plasma drug levels of SOF, its inactive metabolite GS-331007, and DCV were measured at baseline, at day 14 and at EOT (day 28 or 56) in all participants. In addition, intensive drug level sampling was conducted in a subset of 40 participants, who were sequentially invited to join an ancillary PK study. In this subgroup, five samples were collected in each participant after the first dose of SOF/DCV and at day 28, according to one of two randomly assigned sampling schedules (A and B). In sampling schedule A, drug levels were measured at 0.5, 2, 4, 6, and 24 hours post-dose; in sampling schedule B, drug levels were measured at 1, 3, 5, 8 and 24 hours post-dose.

Drug quantification was performed using liquid chromatography tandem mass-spectrometer at Mahidol Oxford Tropical Medicine Research Unit, Bangkok. Two separate analytical assays were developed and validated to quantify SOF plus its metabolite GS-331007, and DCV, respectively. Full methodological details of PK/PD analysis are provided in appendix 1.

### Statistical analysis

#### Primary and secondary outcomes

Analysis were performed under intention-to-treat (the per-protocol analysis, defined as including all participants taking 90-110% of prescribed treatment, was equivalent to the intention-to-treat analysis) with an additional post-hoc analysis excluding those who were non-G1/6 from WGS. Where possible, proportions and 95% confidence intervals (CIs) were estimated from the marginal effects after logistic regression. Where no events were recorded and models would not converge, we used binomial exact 97.5% CIs. Absolute HCV VL was analysed using interval regression (incorporating censoring at the LLOQ) adjusting for baseline HCV VL. Differences between baseline means and medians in 4-weeks cures vs 4-week failures were analysed with unpaired t-tests and Wilcoxon rank sum tests respectively; differences in proportions were assessed using chi-squared tests or Fisher’s exact tests as appropriate. Analyses were performed using Stata v16.1^27^.

### Virus genomics

Fisher’s exact test was used to test for association between presence and absence of each RAS and treatment outcome. To test for association between outcome and number of RAS we used logistic regression.

### PK/PD

Intensive drug levels of SOF, its metabolite GS-331007, and DCV from the subset of 40 patients at day 0 and day 28, together with any EOT samples at day 28, were analysed using non-compartmental analysis in PKanalix version 2020R1^28^. Two separate analyses were performed to characterise the pharmacokinetic properties of the study drugs.

In the first, naïve pooled analyses were performed separately on data from day 0 and day 28 (not including end of treatment samples) to derive pharmacokinetic parameters at each day. In these analyses, the median concentration at each protocol time were calculated. Individual concentration measurements below the LLOQ was set to LLOQ/2 when calculating the median values. It was assumed that the participants had no drug concentrations at time 0.

In the second analysis, data from day 0 and day 28 were pooled for each individual. This resulted in a full pharmacokinetic profile for each subject, which was analysed with a non-compartmental approach. If patients had samples taken at the same time point the mean of the samples were taken. These derived individual drug exposures were used to evaluate the relationship between drug exposure and therapeutic outcome. It was assumed that the participants had no drug concentrations at time 0. In this analysis the first measurement below LLOQ in a series of LLOQ samples were imputed as LLOQ/2 and the later measurements were ignored. In both approaches, the 24-hour samples for SOF were excluded. SOF has a very short half-life, which make concentrations at 24 hours after dose unlikely.

In addition, outcome varibles and the relationship between outcome variables and drug exposure were evaluated.

Additional detail of the PK/PD analysis is provided in appendix 1.

### Ethical approval

The trial was approved by the research ethics committees of The Hospital for Tropical Diseases^29^, Vietnam Ministry of Health^30^, Imperial College London^31^, and Oxford University Tropical Research Ethics Committee^32^. The study’s conduct and reporting is fully compliant with the World Medical Association’s Declaration of Helsinki on Ethical Principles for Medical Research Involving Human Subjects^33^. The trial was registered at ISRCTN, registration number is ISRCTN17100273^34^.

## RESULTS

### Baseline characteristics

Of 455 patients screened, 52 were enrolled and one subsequently withdrew (figure 2). Most exclusions were on account of a either a fibroscan score of >7.1kPa (with cirrhotic patients enrolled into a parallel study^35^),or ineligible genotype.

**Figure 2:**
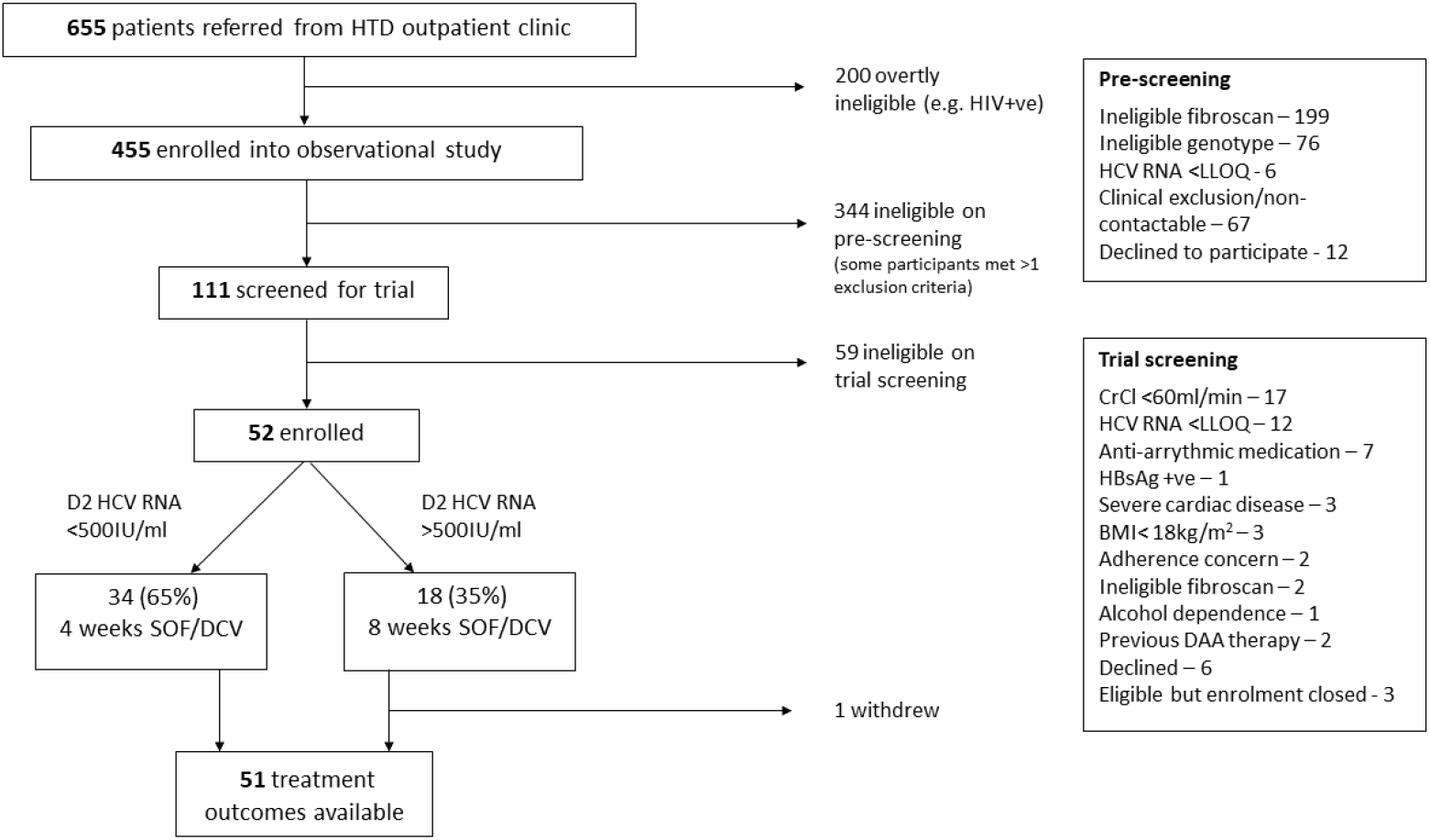
Screening and enrolment.

22/51 were initially identified as genotype 1 infection and 30 as genotype 6. With the benefit of WGS data, it was confirmed that 22 (43%) had genotype 1 infection, 27 (53%) had genotype 6, one had genotype 2 and another had genotype 4 infection. The latter two individuals were included in the intention-to-treat analysis but excluded from a post-hoc analysis of G1 and G6 infections only.

Recruitment was completed short of the initial target of 60 due to severe COVID-19-related restrictions in Vietnam from February 2020. These included clinic closures, travel restrictions and repurposing of the HTD as a COVID-19 treatment centre. Baseline and clinical characteristics are described in Table 1.

**Table 1:** Baseline characteristics.

|  | N/ median | %/range |
| --- | --- | --- |
| Total participants | 52 |  |
| Age in years | 49.5 | (25.0, 67.0) |
| Female | 29 | (56%) |
| Body-mass index in kg/m<sup>2</sup> | 23.3 | (18.7, 30.6) |
| Genotype 1 | 22 | (43%) |
| 1a | 11 |  |
| 1b | 12 (1 withdrew) |  |
| Genotype 6 | 27 | (53%) |
| 6a | 12 |  |
| 6e | 10 |  |
| 6h | 2 |  |
| 6l | 2 |  |
| 6u | 1 |  |
| Genotype 2(m) | 1 |  |
| Genotype 4(k) | 1 |  |
| Baseline HCV viral load in IU/ml | 1,932,775 | (618, 11,200,000) |
| HCV viral load – log<sub>10</sub> IU/ml (range) | 6.3 | (2.8, 7.0) |
| **Past medical history:** |  |  |
| Illicit drug use | 4 | (8%) |
| Alcohol dependence (historic; current excluded) | 4 | (8%) |
| Diabetes | 2 | (4%) |
| Hypertension | 7 | (13%) |
| Ischaemic heart disease | 1 | (2%) |
| Tuberculosis | 2 | (4%) |
| Current smoker | 18 | (35%) |
| Previous spontaneous clearance of HCV with re-infection | 2 | (4%) |

### Treatment duration, adherence and efficacy outcomes

By day two, 34 participants (65%) had HCV viral load below the threshold of 500 IU/ml (figure 2; table 2), so received 4 weeks of treatment. 18 participants were above the threshold at this timepoint, of which one withdrew after 9 days of treatment, meaning 17 completed 8 weeks therapy. Adherence was good, with 96% completing the full prescribed course of SOF/DCV. 18 (35%) participants missed at least one visit because of COVID-19-related restrictions.

**Table 2:** treatment outcome.

|  | N/ median | %/range |
| --- | --- | --- |
| Detectable HCV viral load (HCV VL) at day 2 | 50 | 96% |
| Median (IQR) HCV VL at day 2 in IU/ml | 269 | (104, 690) |
| Below threshold - for 4 weeks therapy | 34 | (65%) |
| Above threshold – for 8 weeks therapy | 18 | (35%) |
| Mean (SD) duration of first-line therapy received in days | 37 | (13.7) |
| Mean (SD) duration of all therapy received in days | 58 | (34.2) |
| Median weeks from enrolment to last visit (range) | 20 | (1, 42) |
| **Primary Outcome** |  |  |
| Outcome available | 51 |  |
| SVR12 by intention-to-treat analysis and per protocol analysis | 38 | (75% [95% CI 63, 86]) |
| SVR12 by sensitivity analysis (i) [missing results = failure] | 38 | (73% [95% CI 61, 85]) |
| SVR12 by post-hoc analysis (ii) [G1 and G6 only] | 37 | (76% [95% CI 63, 88]) |
| **Secondary Endpoints** |  |  |
| Lack of initial virological response | 0 | (0% [97.5% CI 0, 7])* |
| Serious adverse events | 0 | (0% [97.5% CI 0, 7])* |
| Grade 3/4 clinical adverse events | 0 | (0% [97.5% CI 0, 7])* |
| Non-serious adverse reactions | 18 | (35% [95% CI 22, 48]) |
| Adverse events or reactions leading to change in study medication | 0 | (0% [97.5% CI 0, 7])* |

Of the 51 participants with outcome data, 38 (75% [95% CI (63, 86)]) achieved SVR12 while 13 failed therapy and required retreatment. All treatment failures occurred in individuals who received 4 weeks therapy, translating to an SVR12 of 62% (21/34; 95% CI (44, 78)) in rapid responders who received 4 weeks therapy, and 100% (17/17; 97.5% CI (80, 100)) in slower responders who received 8 weeks SOF/DCV (figure 3; table 2).

**Figure 3:**
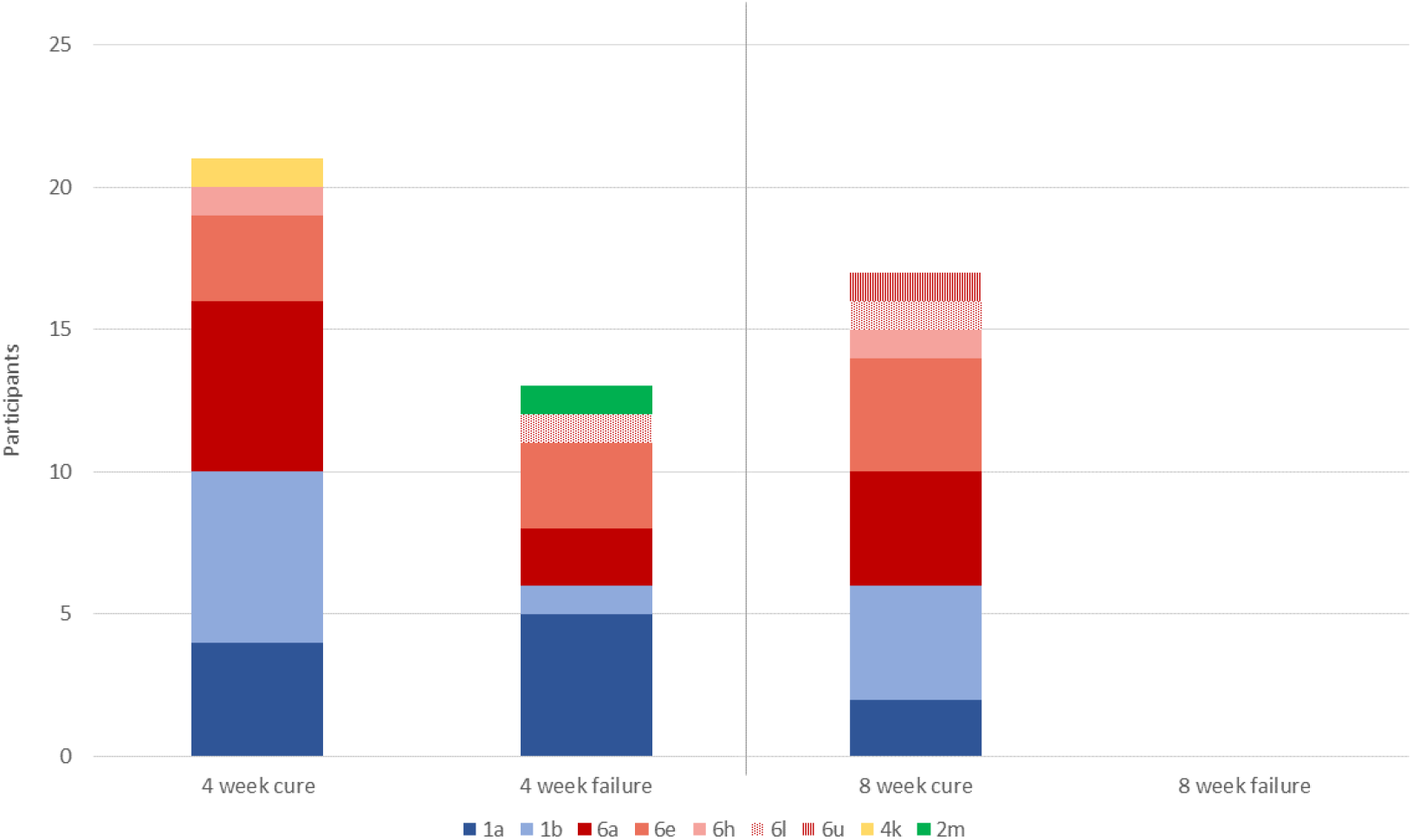
Primary outcome, with HCV subtypes. All 13 individuals who experience treatment failure with 4 weeks SOF/DCV were cured with 12 weeks SOF/DCV retreatment

Of the 13 participants who underwent retreatment, 100% were cured. The mean first-line SOF/DCV treatment duration was 37 days (standard deviation, SD 13.7), with a first-line cure rate of 75%. The mean (SD) total SOF/DCV duration (i.e. including 12 weeks retreatment where required), was 58 (34.2) days per patient, with a 100% cure rate. There was no evidence of differences in age, gender, BMI, IFNL4 genotype, transaminases or baseline HCV viral load between patients who achieved cure with 4-weeks of treatment versus those who experienced treatment failure with 4 weeks of treatment (table 3).

**Table 3:**
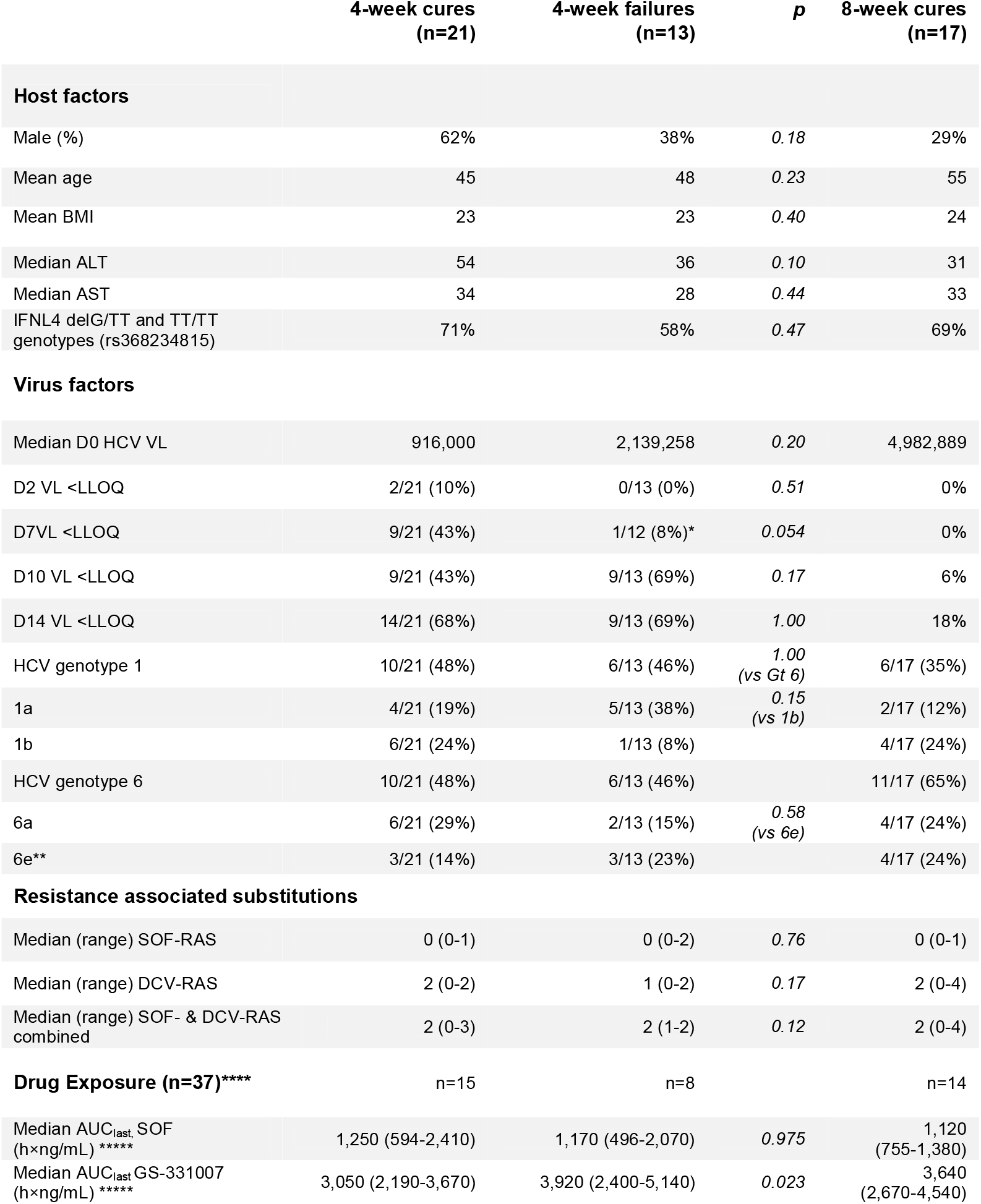
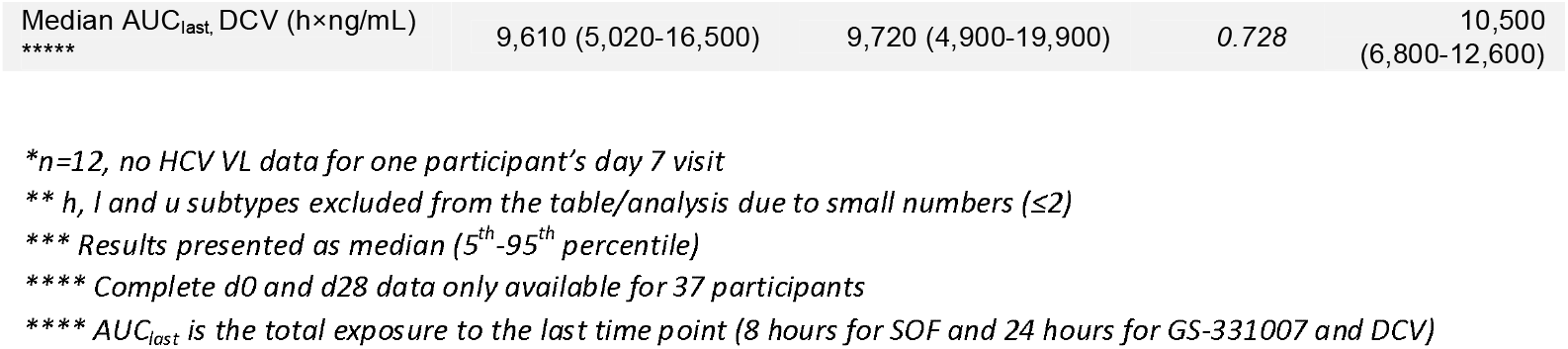
Comparison of baseline factors, drugs levels and virological response in individuals failed to achieve SVR12 with 4 weeks therapy vs those who cured with 4 or 8 weeks therapy.

### Viral kinetics and timing of treatment failure

All participants had an initial virological response ((i.e. ≥1 log10 decrease in HCV viral load from baseline) (appendix 1 - figure 1)). There was no evidence of association between time to complete virological suppression (HCV RNA <LLOQ) and treatment outcome (table 3; appendix 1 – figure 2). In an exploratory analysis, we estimated first-line cure rates based on suppression below the LLOQ at other timepoints which could be used for RGT. At day 7, 9/21 cures and 1/12 treatment failures (one missed visit) had HCV RNA <LLOQ (p=0.054; table 3), translating to 90% sensitivity (95% CI [56, 100]) for predicting cure with 4 weeks treatment. However, by day 10, 9/21 cures and 9/13 failures had HCV RNA <LLOQ (p=1.00), making a rapid virological response 50% [26, 74] sensitive in predicting cure with 4 weeks treatment. This suggests early on-treatment response alone may be of limited value in determining cure with ultra-short therapy.

All treatment failures occurred during follow-up after EOT. Despite intensive twice weekly sampling from EOT to EOT+28d, the earliest virologic rebound occurred 3 weeks after completion of therapy (appendix 1 - figure 3).

### Viral genomics at baseline

Whole genome sequencing was attempted on all participants’ virus at baseline, but consensus sequences could not be assembled in two individuals (who had low baseline viral load and were both cured with first line therapy). This left 50 patients with baseline sequences, of which 49 had outcome data.

We found nine discrepancies between lab genotyping and sequencing-based genotyping. Five of these differences were at the level of subtypes for genotype 6 samples, highlighting difficulties inherent in classifying this rare and genetically diverse lineage using an amplicon approach for genotying (lab genotyping). Two samples were called 6a/e using lab genotyping and whole genome sequencing classified them as 6e. One sample was classified as 6e on lab genotyping, but whole genome sequencing showed that it was a genotype 2m sample. Whole genome sequencing revealed another patient to have mixed infection with genotype 1a and genotype 6a; this had been classified by laboratory genotyping as a genotype 6a mono-infection. The individual with mixed infection received 4 weeks of SOF/DCV but cure was not achieved, with relapse of the genotype 1a infection. They subsequently responded to 12 weeks retreatment.

We found no evidence of differences between genotypes or subtypes with regards to rates of treatment failure. Among genotype 1-infected individuals, 1/7 subtype 1b infections experienced treatment failure with 4 weeks therapy compared with 4/8 subtype 1a infections (including the mixed infection case) (p=0.15). Among genotype 6-infected individuals, 1/8 subtype 6a infections were not cured with 4 weeks SOF/DCV compared with 3/6 subtype 6e (p=0.58), 0/1 subtype 6h and 1/1 subtype 6l.

At baseline, the 159F SOF RAS was identified in one patient, and the 237G putative SOF RAS was identified in six patients (appendix 1-figures 4 and 5). The DCV RAS 24R, 30R, 31M, 93H and 93S were detected at baseline (appendix 1 - figures 6 and 7).

In the assessment of SOF RAS (appendix 1-figure 5), the one patient who had 159F at baseline failed treatment, although this was a minority variant making up 20% of the sequencing reads (figure 4; black box). 237G was identified as a majority variant in two individuals where treatment failed, but was also seen in four individuals who were cured (three received 4 weeks treatment (appendix 1 - figure 5).

**Figure 4:**
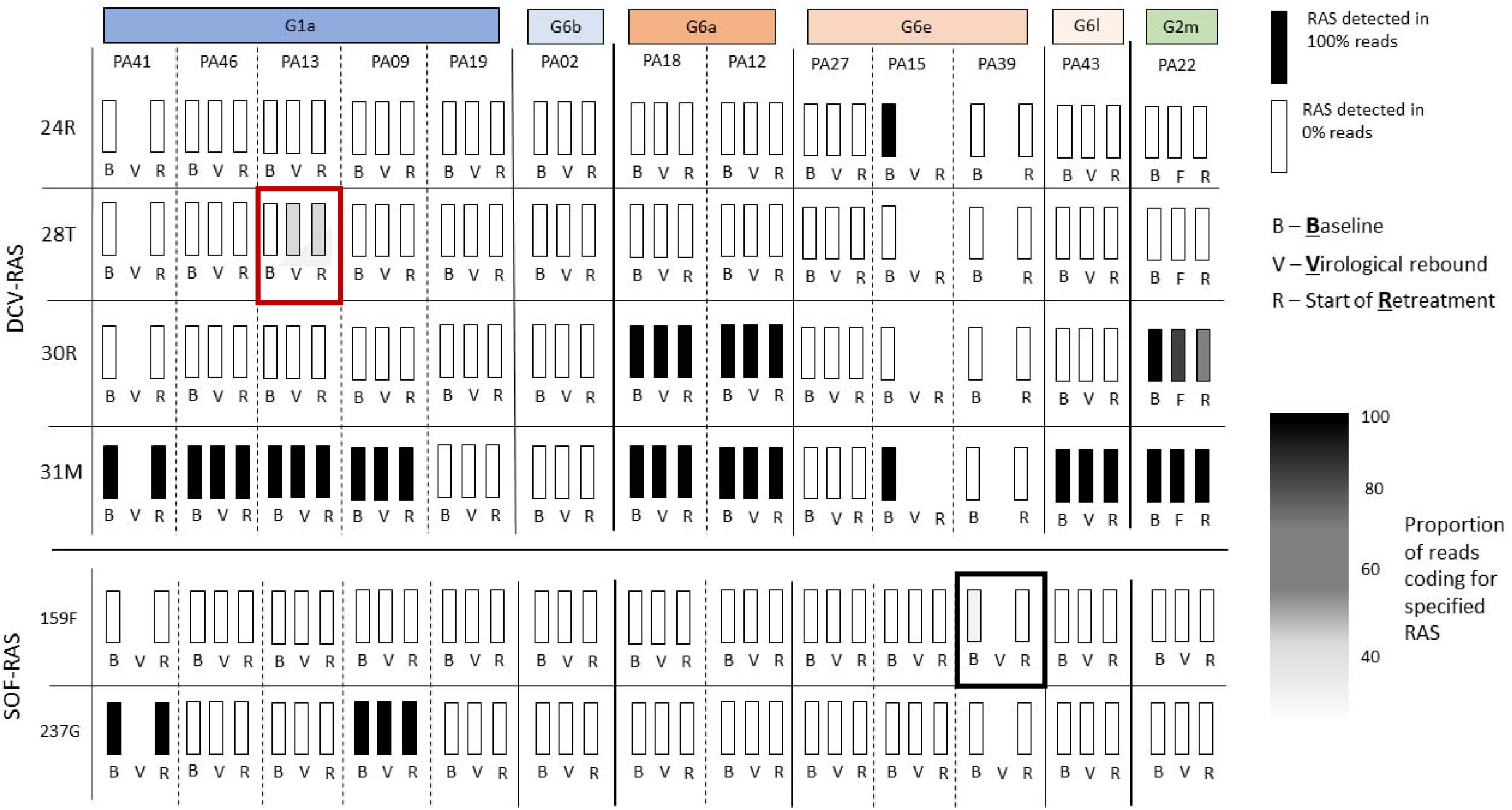
Sofosbuvir RAS and Daclatasvir RAS at baseline, treatment failure, and at start of retreatment in all participants who failed first line treatment.

The most prevalent DCV RAS was 31M, present in 9 participants where treatment failed after 4 week first-line therapy (figure 4; appendix 1 - figures 8 and 9). However, 31M was also found in 13 individuals cured with 4 weeks treatment, and 13 cured with 8 weeks. The next most prevalent RAS was 30R, present at baseline in 3 patients who had treatment failure, in 5 individuals cured with 4 weeks treatment and in 4 patients cured with 8 weeks treatment. 30R RAS was present in 11/12 6a genomes and 1/1 2m genomes but was absent in other subtypes. 31M RAS was present in 10/11 1a genomes and 12/12 6a genomes and was also found in other subtypes (appendix 1 – figures 7 and 8). Additionally almost all of the subtype 6a samples carried both 30R and 31M RASs while other subtypes did not carry this combination (apart from the 2m sample).

### Viral genomics in participants failing first line therapy

Among 13 individuals who experienced treatment failure, we compared the emerging viral genome with baseline virus (figure 4). Full genome sequences could not be assembled for three participants at time of virological relapse, however, we were able to generate whole viral genomes using samples from the start of retreatment for two of these individuals. No new genomes were identified at treatment failure (ruling out any new infections). No new SOF RAS were identified on virologic rebound. DCV 28T RAS (not present at baseline) was identified in one participant failing therapy (see red box figure 4; figure S9) as a minority variant at time of virological rebound and start of retreatment (at 30% and 25% of reads respectively). Given 100% of retreated individuals achieved SVR12 with standard duration of therapy we found no evidence to suggest this emerging RAS was clinically significant. There was no evidence of differences in the number of combined SOF- and DCV-RAS at baseline in those who failed 4 week therapy (median 2, range 0-3) vs those who cured with 4 weeks (median 2, range 1-2), (p=0.12) or in those with a slower initial virological response, who received 8 weeks (median 2, range 0-4).

### Pharmacokinetics and pharmacodynamics

Pharmacokinetic parameters derived from the naïve pooled analysis (based on 40 patients on day 0 and 37 and day 28) are presented in appendix 1 - table 1. Exposure after the individual analysis as well as outcome measurements are presented in appendix 1 - table 2. In the individual analysis and the linear regression between outcome measurements and drug exposure 3 patients were excluded as they did not have dense samples collected at day 28 (n=37). In the analysis of outcome variables data from all 40 patients were used. No significant relationship between outcome variables and drug exposure was found using linear regression (appendix 1 - table 3).

In the subset of 37 participants who underwent dense PK analysis at d0 and 28, 23 received 4 weeks SOF/DCV and 14 received 8 weeks. There was no significant difference between AUC_last_ (total drug exposure to the last time point) for SOF and DCV in 4 week cures (n=15) vs 4 week failures (n=8); (table 3). However GS-331007 levels were significantly higer in treatment failures (p=0.032).

### Safety

SOF/DCV was well-tolerated in general and no participants discontinued treatment due to drug side effects. 18 participants (35%; 95% CI 22%, 48%) reported at least one non-serious adverse reaction. The most common of these were insomnia, gastritis and dizziness, which are all consistent with undesirable effects described in the summary of product characteristics of SOF/DCV^36^. There were no serious adverse events or grade 3 or 4 adverse events.

## DISCUSSION

In this mechanistic study in individuals with genotype 1 or 6 HCV infection and mild liver disease, treated with 4 or 8 weeks of SOF/DCV depending on HCV viral load 2 days after starting treatment, overall first-line cure rate was 75% [95% CI (63, 86)], with a mean 37 days treatment. This saved 47 days DAA therapy per participant compared with a standard 12 week course, but cure rate fell below our target of ≥90%. For the secondary endpoint - SVR12 after combined first-line therapy or retreatment - cure was 100%, with mean treatment duration 58 days, saving 26 days DAAs per participant.

### Effect of shortening therapy

Inferior rates of cure are well described when DAA therapy is shortened below 8 weeks without use of early on-treatment virological response, falling below 50% with ≤4 weeks therapy without stratification^7,37,38^. A few small studies have reported high rates of cure with shortened therapy based early virological response^8,39,40^. The only previous RGT study to use less than 6 weeks treatment, by Lau et el, found a cure rate of 100% with just three weeks of DAA therapy in 18 individuals whose HCV viral load was suppressed below 500 IU/ml after two days of therapy. This was the same threshold and time point used in our study. One important difference was in the treatment regimen, which included a protease inhibitor (simeprevir or asunaprevir). Although NS5A-(DCV) and NS5B-(SOF) inhibitors rapidly eliminate HCV from the blood, second-phase decline in viral load appears to be enhanced by addition of a protease inhibitor^41^. This may be crucial in sustaining high rates of cure with shortened therapy. Additionally, 100% of participants in that study had genotype 1b infection, compared with just 23% (n=12) in ours. Genotype 1b is associated with favourable outcomes with some DAAs^42,43^. Although real world 1b outcomes with standard duration SOF/DCV appear similar to other non-3 genotypes^44^, our data are consistent with the hypothesis that subtype may become more important when treatment is shortened.

### Role for response-guided therapy with SOF/DCV

Cure rates with this strategy are too low for it to be routinely recommended. With standard duration therapy, SVR12 is known not to be impacted by time to first undetectable HCV RNA^45^ or by the presence of detectable virus at the end of treatment ^46^. This also appears to be true of shortened treatment: in one individual who experienced treatment failure, HCV viral load was already <LLOQ by day 7; in five of the 4-week cures, HCV VL was only suppressed to <LLOQ virus for the first time at end of treatment (appendix 1 - figure 2). Comparison of 4-week cures and 4-week treatment failures inidicates that an HCV RNA <LLOQ by day 7 may be a useful discriminator of 4-week treatment outcome (p=0.054). However, day 10 HCV RNA<LLOQ was not predictive of response to shortened treatment. Day-7 viral load thresholds for shortening DAA therapy are currently being evaluated as part of a large ongoing randomised controlled trial in Vietnam^47^.

### A case for 8-weeks SOF/DCV therapy

Given the high rates of cure observed with 8 weeks of SOF/DCV in participants with a slow initial virological response (17/17), there is a case for reducing SOF/DCV therapy from 12 to 8 weeks in individuals with mild liver disease. Pror evidence for caution regarding 8 weeks of SOF/DCV comes predominantly from a small 2015 study in HIV-coinfected individuals^48^, in which 7/10 treatment failures in the 8-week arm received half-dose daclatasvir (30mg) because participants were taking concomitant darunavir–ritonavir. This dose adjustment was subsequently deemed unnecessary once drug-interaction data emerged, such that this study is likely to underestimate the efficacy of 8 weeks SOF/DCV. More recent studies corroborate our finding of >90% cure with 8 weeks NS5A/NS5A inhibitor combination^39,49,50^. Larger trials are warranted to evaluate 8 weeks SOF/DCV therapy for patients with mild liver disease (irrespective of speed of virological response). This could save significant costs, particularly in countries where pricing is determined per pill rather than per treatment course, such as Vietnam, and the USA^37,51^.

### Impact of resistance-associated substitutions and retreatment concerns

To our knowledge this study is the largest assessment of G6 RAS in vivo with SOF/DCV therapy. We hypothesised that a high number of putative RAS at baseline may be associated with higher rates of failure with shortened treatment. However, we found no evidence that number or type of SOF- or DCV-RAS was different at baseline in 4-week cures compared with 4-week treatment failures (table 3, appendix 1 - figures 5 and 7), although numbers were small. Additionnally, the excellent retreatment outcomes observed (13/13) are reassuring, particularly for low-resource settings where protease inhibitor-based retreatment options are limited. Only one novel RAS was detected after first line treatment failure, and the individual concerned achieved SVR with standard duration retreatment, suggesting this was not clinically relevant.

### Impact of drug levels

This was the first assessment of the impact of DAA drug levels on efficacy of shortened therapy. The inactive SOF metabolite GS-331007 is the main circulating metabolite of SOF prior to undergoing renal excretion, and it is frequently used to describe SOF’s pharmacokinetics^52^. We hypothesized that accumulation and slow elimination of GS-331007 and DCV in vivo might protect against the re-emergence of HCV viraemia. However we found no evidence of a difference in AUC_last_ between 4-week cures and 4-week treatment failures for SOF or DCV. Total exposure to GS-331007 was higher in treatment failures (3,920 (2,400-5,140) vs 3,050 (2,190-3,670) (p=0.023). This was a surprising result, given that SOF and GS-331007 AUCs are near dose proportional over the dose range of 200 mg to 1200 mg^52^, and higher day 10 concentrations of GS-331007 have been associated with improved rates of cure with SOF/ribavirin treatment^53^. Further PK studies are warranted to better understand if SOF metabolism impacts treatment outcomes.

### Limitations

Our study has important limitations. Firstly it was powered to determine overall cure rate with 4- and 8-weeks treatment, rather than outcomes with each duration. It is possible that we would have seen patients failing 8 weeks therapy with a larger sample, and our cure estimates may therefore be imprecise. Secondly, the participating cohort did not include any individuals with HIV, Hepatitis B-co-infection or renal impairment and only 4 participants reported a history of injecting drug use, of which none were currently injecting. These populations are known to have an altered immunological response and constitute an important part of the HCV epidemic. Thirdly, in order to identify the timing of failure, the protocol required a visit schedule with many more visits than is standard of care, which many patients would not be able to follow. Consequently, adherence was very high, which may not reflect real world practice. Finally our non-compartmental analysis of drug levels may not adequately account for drug accumulation of sofosbuvir’s metabolite GS-331007 and DCV between day 0 and 28, which was observed (see appendix 1 for more detail).

In summary our findings indicate that shortened SOF/DCV therapy cures a significant proportion of patients with mild liver disease without compromising retreatment with the same drug combination in those who fail first-line therapy. This study adds to a growing case for shortening SOF/DCV therapy in individuals with mild liver disease from 12 to 8 weeks, and offering retreatment with 12 weeks SOF/DCV when required. There was no evidence that relatively high numbers of putative resistance associated substitutions at baseline were associated with treatment outcomes, suggesting routine sequencing at baseline or prior to retreatment remains unnecessary. We also found no evidence that drug levels affect virological response or influence treatment outcome. Further work is required to understand which factors predict cure with ultra-short DAA treatment.

## Data Availability

TThe study protocol and processed study data have been uploaded to the ISRCTN registry (ISRCTN17100273; https://doi.org/10.1186/ISRCTN17100273). The data are available under unrestricted access. The raw data are protected and are not available due to data privacy laws. The virus sequencing dataset has been uploaded to Dryad (https://datadryad.org) and is available here: doi:10.5061/dryad.x0k6djhnp. All data generated in this study is provided in the main text or appendix 1.

https://doi.org/10.5061/dryad.x0k6djhnp

https://doi.org/10.1186/ISRCTN17100273

## Acknowledgements

This work was supported by the Medical Research Council (grant MR/P025064/1) and The Global Challenges Research Fund (Wellcome Trust Grant 206/296/Z/17/Z). GC is supported in part by the NIHR Biomedical Research Centre of Imperial College NHS Trust and an NIHR Professorship. BF received a travel grant to attend AASLD conference in Washington DC from Gilead Sciences in 2017. HCT acknowledges funding from the MRC Centre for Global Infectious Disease Analysis (reference MR/R015600/1), jointly funded by the UK Medical Research Council (MRC) and the UK Foreign, Commonwealth & Development Office (FCDO), under the MRC/FCDO Concordat agreement and is also part of the EDCTP2 programme supported by the European Union. EB acknowledges support from Oxford NIHR Biomedical Research Centre. MAA is supported by a Sir Henry Dale Fellowship jointly funded by the Royal Society and Wellcome Trust (220171/Z/20/Z). ASW and EB are NIHR Senior Investigators. LM, SLP and ASW are supported by core support from the Medical Research Council UK to the MRC Clinical Trials Unit [MC_UU_00004/03]. The views expressed are those of the author(s) and not necessarily those of the NIHR or the Department of Health and Social Care. No authors report conflicts of interest relating to this work.

We would like to thank the patients of the HTD who volunteered to participate in the trial, our hard-working nurses An Nguyen Thi Chau, Tan Dinh Thi, Nga Tran Thi Tuyet, and Phuc Le Thi, as well as members of the data monitoring committee: Hoa Pham Le, Timothy Peto, and John Dillon.

## Data Availability

The study protocol and processed study data have been uploaded to the ISRCTN registry (ISRCTN17100273; https://doi.org/10.1186/ISRCTN17100273). The data are available under unrestricted access. The raw data are protected and are not available due to data privacy laws. The virus sequencing dataset has been uploaded to Dryad (https://datadryad.org) and is available here: doi:10.5061/dryad.x0k6djhnp. All data generated in this study is provided in the main text or appendix 1.

---

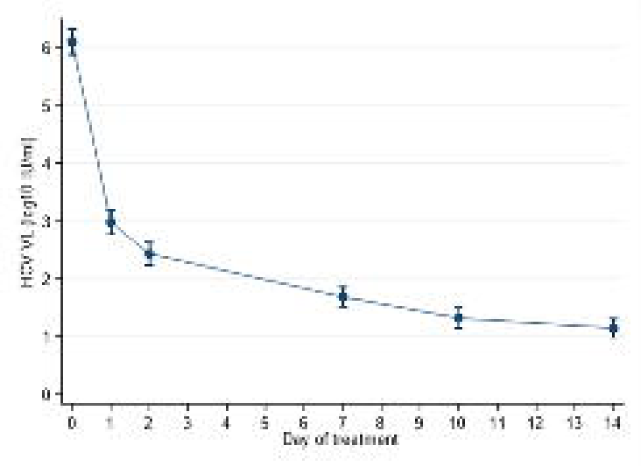

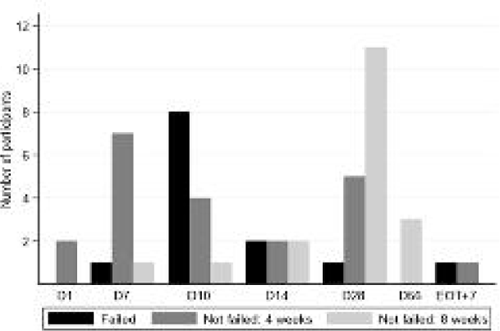

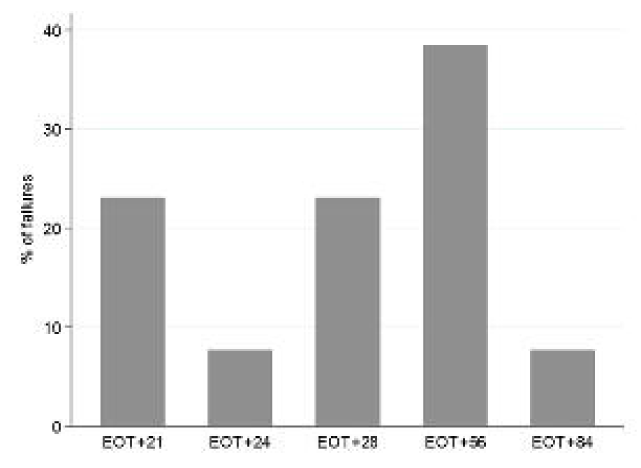

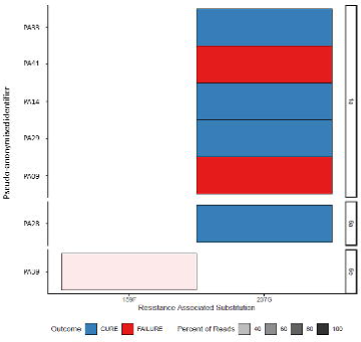

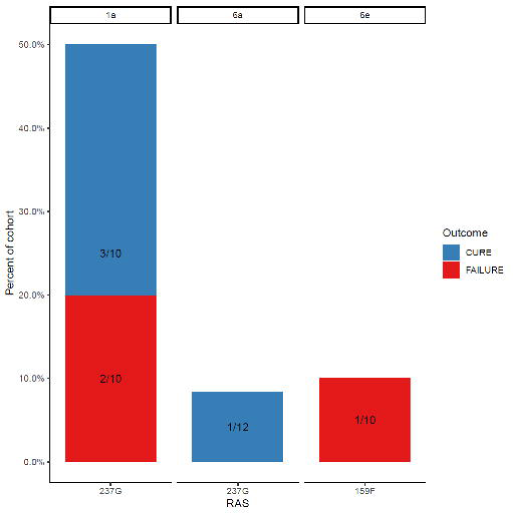

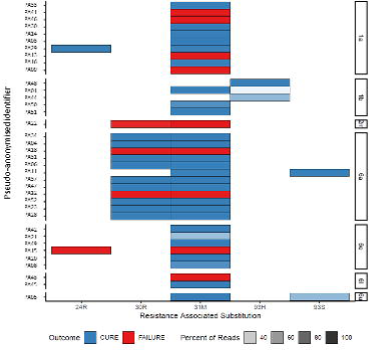

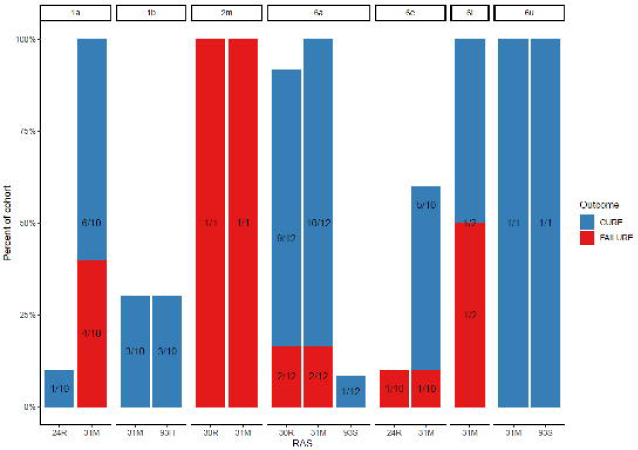

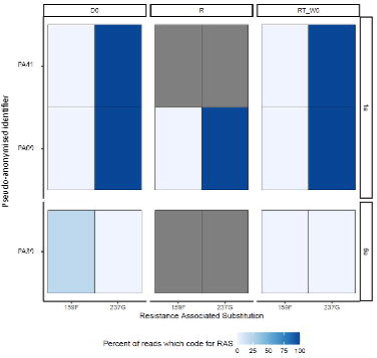

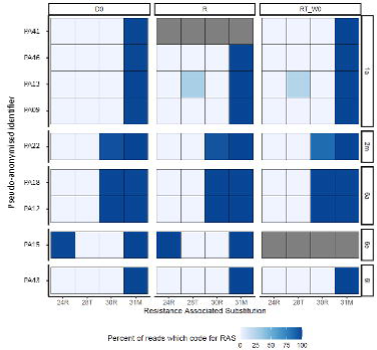

## REFERENCES

1. Kracht PAM, de Gee EA, van der Poel A, et al. Introducing hepatitis C virus healthcare pathways in addiction care in the Netherlands with a Breakthrough project: a mixed method study. Harm Reduct J. 2019;16(1):45. doi:10.1186/s12954-019-0316-4

2. Petersen T, Townsend K, Gordon LA, et al. High adherence to all-oral directly acting antiviral HCV therapy among an inner-city patient population in a phase 2a study. Hepatol Int. 2016;10(2):310–319. doi:10.1007/s12072-015-9680-7

3. Nguyen Thanh D, Tran Thi Thanh T, Nghiem My N, et al. Baseline Characteristics and Treatment Cost of Hepatitis C atHospital for Tropical Diseases, Ho Chi Minh City, Vietnam inDirect-Acting Antiviral Treatment Era. J Gastroenterol Hepatol Res. 2019;8(5):2962–2971. doi:10.17554/j.issn.2224-3992.2019.08.857

4. Ministry of Health V. National Plan for Prevention and Control of Viral Hepatitis, Period 2021-2025. 2021. https://emoh.moh.gov.vn/publish/attach/getfile/408692.

5. World Health Organization (WHO). Guidelines for the Care and Treatment of Persons Diagnosed with Chronic Hepatitis C Virus Infection.; 2018.

6. Cooke GS, Pett S ML et al. Strategic treatment optimization for HCV (STOPHCV1): a randomised controlled trial of ultrashort duration therapy for chronic hepatitis C [version 2; peer review: 2 approved]. Wellcome Open Res. 2021;6:93. https://doi.org/10.12688/wellcomeopenres.16594.2.

7. Jones CR, Flower BF, Barber E, Simmons B, Cooke GS. Treatment optimisation for hepatitis C in the era of combination direct-acting antiviral therapy: A systematic review and meta-analysis [version 1; peer review: 2 approved]. Wellcome Open Res. 2019;4. doi:10.12688/wellcomeopenres.15411.1

8. Lau G, Benhamou Y, Chen G, et al. Efficacy and safety of 3-week response-guided triple direct-acting antiviral therapy for chronic hepatitis C infection: a phase 2, open-label, proof-of-concept study. lancet Gastroenterol Hepatol. 2016;1(2):97–104. doi:10.1016/S2468-1253(16)30015-2

9. Clinton Health Access Initiative. Hepatitis C Market Report.; 2020.

10. da Silva Filipe A, Sreenu V, Hughes J, et al. Response to DAA therapy in the NHS England Early Access Programme for rare HCV subtypes from low and middle income countries. J Hepatol. 2017;67(6):1348–1350. doi:10.1016/j.jhep.2017.06.035

11. Gupta N, Mbituyumuremyi A, Kabahizi J, et al. Treatment of chronic hepatitis C virus infection in Rwanda with ledipasvir–sofosbuvir (SHARED): a single-arm trial. Lancet Gastroenterol Hepatol. 2019;4(2):119–126. doi:10.1016/S2468-1253(18)30382-0

12. Irekeola AA, Malek NA, Wada Y, Mustaffa N, Muhamad NI, Shueb RH. Prevalence of HCV genotypes and subtypes in Southeast Asia: A systematic review and meta-analysis. PLoS One. 2021;16(5):e0251673. https://doi.org/10.1371/journal.pone.0251673.

13. Hedskog C, Parhy B, Chang S, et al. Identification of 19 novel hepatitis C virus subtypes-further expanding HCV classification. Open Forum Infect Dis. 2019;6(3):1–9. doi:10.1093/ofid/ofz076

14. Fiona McPhee, a* Joseph Ueland, a Vincent Vellucci, a Scott Bowden, b William Sievert c NZ. Impact of Preexisting Hepatitis C Virus Genotype 6 NS3, NS5A, and NS5B Polymorphisms on the In Vitro Potency of Direct-Acting Antiviral Agents. Antimicrob Agents Chemother. 2019;(November 2018):1–12.

15. Prokunina-Olsson L, Muchmore B, Tang W, et al. A variant upstream of IFNL3 (IL28B) creating a new interferon gene IFNL4 is associated with impaired clearance of hepatitis C virus. Nat Genet. 2013;45(2):164–171. doi:10.1038/ng.2521

16. Franco S, Aparicio E, Parera M, Clotet B, Tural C, Martinez MA. IFNL4 ss469415590 variant is a better predictor than rs12979860 of pegylated interferon-alpha/ribavirin therapy failure in hepatitis C virus/HIV-1 coinfected patients. AIDS. 2014;28(1). https://journals.lww.com/aidsonline/Fulltext/2014/01020/IFNL4_ss469415590_variant_is_a_better_predictor.17.aspx.

17. Ansari MA, Pedergnana V, L C Ip C, et al. Genome-to-genome analysis highlights the effect of the human innate and adaptive immune systems on the hepatitis C virus. Nat Genet. 2017;49(5):666–673. doi:10.1038/ng.3835

18. Ansari MA, Aranday-Cortes E, Ip CL, et al. Interferon lambda 4 impacts the genetic diversity of hepatitis C virus. Elife. 2019;8:e42463. doi:10.7554/eLife.42463

19. Nitta Y, Kawabe N, Hashimoto S, et al. Liver stiffness measured by transient elastography correlates with fibrosis area in liver biopsy in patients with chronic hepatitis C. Hepatol Res. 2009;39(7):675–684. doi:https://doi.org/10.1111/j.1872-034X.2009.00500.x

20. National Institutes of Health. Common Terminology Criteria for Adverse Events (CTCAE).; 2017. https://ctep.cancer.gov/protocolDevelopment/electronic_applications/docs/CTCAE_v5_Quick_Reference_8.5x11.pdf.

21. Le Ngoc C, Tran Thi Thanh T, Tran Thi Lan P, et al. Differential prevalence and geographic distribution of hepatitis C virus genotypes in acute and chronic hepatitis C patients in Vietnam. PLoS One. 2019;14(3):e0212734. doi:10.1371/journal.pone.0212734

22. Thomson E, Ip CLC, Badhan A, et al. Comparison of Next-Generation Sequencing Technologies for Comprehensive Assessment of Full-Length Hepatitis C Viral Genomes. Loeffelholz MJ, ed. J Clin Microbiol. 2016;54(10):2470 LP – 2484. doi:10.1128/JCM.00330-16

23. Smith DA, Fernandez-Antunez C, Magri A, et al. Viral genome wide association study identifies novel hepatitis C virus polymorphisms associated with sofosbuvir treatment failure. Nat Commun. 2021;12(1):6105. doi:10.1038/s41467-021-25649-6

24. Smith DA, Bradshaw D, Mbisa JL, et al. Real world SOF/VEL/VOX retreatment outcomes and viral resistance analysis for HCV patients with prior failure to DAA therapy. J Viral Hepat. 2021;28(9):1256–1264. doi:https://doi.org/10.1111/jvh.13549

25. Manso CF, Bibby DF, Lythgow K, et al. Technical Validation of a Hepatitis C Virus Whole Genome Sequencing Assay for Detection of Genotype and Antiviral Resistance in the Clinical Pathway. Front Microbiol. 2020;11. https://www.frontiersin.org/articles/10.3389/fmicb.2020.576572.

26. Bradshaw D, Mbisa JL, Geretti AM, et al. Consensus recommendations for resistance testing in the management of chronic hepatitis C virus infection: Public Health England HCV Resistance Group. J Infect. 2019;79(6):503–512. doi:10.1016/j.jinf.2019.10.007

27. StataCorp. Stata Statistical Software: Release 16. College Station, TX: StataCorp LLC. 2019.

28. Sas L. PKanalix version 2020R1. Antony, France. 2022.

29. Hospital for Tropical Diseases Ethics Commitee V. HTD EC (764 Vo Van Kiet, ward 1, district 5, HCMC, Viet Nam; (+84) 28 3923 8704), ref: CS/BND/18/25 Approved 07/06/ 2018. Presented at the:

30. Vietnam Ministry of Health. Ethical Evaluation Commitee in Biomedical Research of Ministry of Health, Vietnam (138A Giang Vo, Ba Dinh, Ha Noi, Viet Nam; (+84) 0462732156), ref: 6172/QĐ-BYTtnam MoH Ethics. Presented at the:

31. ICREC. Imperial College Research Ethics Committee, UK (Level 2, Medical School Building, St Marys Campus, London W2 1PG; +44 (0)207 594 9484), ref: 17IC4238. Approved 18/12/2018. Presented at the:

32. OXTREC. Oxford Tropical Research Ethics Committee, UK (University of Oxford Research Services, University Offices, Wellington Square, Oxford OX1 2JD; +44 (0) 1865 (2)82106; ref: 43-17 Approved 04/10/2018. Presented at the:

33. World Medical Association. WMA Declaration of Helsinki – Ethical Principles for Medical Research Involving Human Subjects. https://www.wma.net/policies-post/wma-declaration-of-helsinki-ethical-principles-for-medical-research-involving-human-subjects/.

34. Cooke GS. ISRCTN registry. ISRCTN17100273. https://doi.org/10.1186/ISRCTN17100273. Published 2018.

35. Flower B, McCabe L, Le Ngoc C, et al. High cure rates for HCV genotype 6 in advanced liver fibrosis with 12 weeks sofosbuvir and daclatasvir: The Vietnam SEARCH Study. Open Forum Infect Dis. June 2021. doi:10.1093/ofid/ofab267

36. EMA. Daklinza (SOF/DCV) Summary of Product Characteristics. Eur Med Agency. 2014.

37. Emmanuel B, Wilson EM, O’Brien TR, Kottilil S, Lau G. Shortening the duration of therapy for chronic hepatitis C infection. lancet Gastroenterol Hepatol. 2017;2(11):832–836. doi:10.1016/S2468-1253(17)30053-5

38. Cooke GS, Pett S ML et al. Strategic treatment optimization for HCV (STOPHCV1): a randomised controlled trial of ultrashort duration therapy for chronic hepatitis C. [version 1; peer Rev 1 Approv 1 Approv with Reserv Wellcome Open Res /wellcomeopenres165941). 2021;6:93. doi:https://doi.org/10.12688

39. El-Shabrawi MH, Abdo AM, El-Khayat HR, Yakoot M. Shortened 8 Weeks Course of Dual Sofosbuvir/Daclatasvir Therapy in Adolescent Patients, With Chronic Hepatitis C Infection. J Pediatr Gastroenterol Nutr. 2018;66(3). https://journals.lww.com/jpgn/Fulltext/2018/03000/Shortened_8_Weeks_Course_of_Dual.15.aspx.

40. Etzion O, Dahari H, Yardeni D, et al. Response guided therapy for reducing duration of direct acting antivirals in chronic hepatitis C infected patients: a Pilot study. Sci Rep. 2020;10(1):17820. doi:10.1038/s41598-020-74568-x

41. Perelson AS, Guedj J. Modelling hepatitis C therapy--predicting effects of treatment. Nat Rev Gastroenterol Hepatol. 2015;12(8):437–445. doi:10.1038/nrgastro.2015.97

42. Kohli A, Osinusi A, Sims Z, et al. Virological response after 6 week triple-drug regimens for hepatitis C: a proof-of-concept phase 2A cohort study. Lancet (London, England). 2015;385(9973):1107–1113. doi:10.1016/S0140-6736(14)61228-9

43. Ferenci P, Bernstein D, Lalezari J, et al. ABT-450/r–Ombitasvir and Dasabuvir with or without Ribavirin for HCV. N Engl J Med. 2014;370(21):1983–1992. doi:10.1056/NEJMoa1402338

44. Charatcharoenwitthaya P, Wongpaitoon V, Komolmit P, et al. Real-world effectiveness and safety of sofosbuvir and nonstructural protein 5A inhibitors for chronic hepatitis C genotype 1, 2, 3, 4, or 6: a multicentre cohort study. BMC Gastroenterol. 2020;20(1):47. doi:10.1186/s12876-020-01196-0

45. Kowdley K V, Nelson DR, Lalezari JP, et al. On-treatment HCV RNA as a predictor of sustained virological response in HCV genotype 3-infected patients treated with daclatasvir and sofosbuvir. Liver Int Off J Int Assoc Study Liver. 2016;36(11):1611–1618. doi:10.1111/liv.13165

46. Pal V, Ancha N, Mann J, Modi AA. Effect of Low Positive End of Treatment Viral Load with Direct-Acting Antiviral Therapy on Sustained Virologic Response. Peltekian KM, ed. Can J Gastroenterol Hepatol. 2020;2020:8815829. doi:10.1155/2020/8815829

47. McCabe L, White IR, Chau NVV, et al. The design and statistical aspects of VIETNARMS: a strategic post-licensing trial of multiple oral direct-acting antiviral hepatitis C treatment strategies in Vietnam. Trials. 2020;21(1):413. doi:10.1186/s13063-020-04350-x

48. Wyles DL, Ruane PJ, Sulkowski MS, et al. Daclatasvir plus Sofosbuvir for HCV in Patients Coinfected with HIV-1. N Engl J Med. 2015;373(8):714–725. doi:10.1056/NEJMoa1503153

49. Yakoot M, Abdo AM, Abdel-Rehim S, Helmy S. Response Tailored Protocol Versus the Fixed 12Weeks Course of Dual Sofosbuvir/Daclatasvir Treatment in Egyptian Patients With Chronic Hepatitis C Genotype-4 Infection: A Randomized, Open-label, Non-inferiority Trial. EBioMedicine. 2017;21:182–187. doi:10.1016/j.ebiom.2017.05.011

50. Boyle A, Marra F, Peters E, et al. Eight weeks of sofosbuvir/velpatasvir for genotype 3 hepatitis C in previously untreated patients with significant (F2/3) fibrosis. J Viral Hepat. 2020;27(4):371–375. doi:10.1111/jvh.13239

51. Clinton Health Access Initiative. HCV Market Intelligence Report 2021 and Preliminary HBV Market Insights. 2021:23–34.

52. Smolders EJ, Jansen AME, ter Horst PGJ, Rockstroh J, Back DJ, Burger DM. Viral Hepatitis C Therapy: Pharmacokinetic and Pharmacodynamic Considerations: A 2019 Update. Clin Pharmacokinet. 2019;58(10):1237–1263. doi:10.1007/s40262-019-00774-0

53. Ahmed B, Munir B, Ghaffar A, et al. Pharmacokinetics profile of serum and cellular Sofosbuvir along with its concentration effect analysis in HCV patients receiving Sofosbuvir and Ribavirin. Pak J Pharm Sci. 2019;32.

