## Appendix for "Efficacy of ultra-short, response-guided sofosbuvir and daclatasvir therapy for Hepatitis C: a single arm mechanistic pilot study"

**APPENDIX 1**

**Efficacy of ultra-short, response-guided sofosbuvir and daclatasvir therapy for Hepatitis C: a single arm pilot mechanistic study**

**Pharmacokinetic and Pharmacodynamic (PK/PD) process**

Sofosbuvir and GS-331007 were extracted from 100 µL of plasma using phospholipid removal in the 96-well plate format (Phree, 8E-S133-TGB, Phenomenex), followed by separation on a Gemini, 50 mm × 2.0 mm I.D. 5 µm, column (00B-4435-B0, Phenomenex). Quantification was performed using selected reaction monitoring for the transitions m/z 530.2–>243.2 (sofosbuvir), 536.2–>243.1 (isotope-labelled internal standard for sofosbuvir), 261.3–>113.1 (GS-331007), and 265.3–>113.1 (isotope-labelled internal standard for GS-331007).

The lower limit of quantification (LLOQ) was set to 1.95 ng/mL for sofosbuvir and 20.5 ng/mL for GS-331007. A total of 9 quality control samples (3×low, 3×mid and 3×high concentration) were analysed for each analyte within each batch of clinical samples (96-well plate), resulting in an accuracy of 2.81-2.93% RSE for sofosbuvir and 2.19-3.50% RSE for GS-331007.

DCV was extracted from 100 µL of plasma using supported liquid extraction in the 96-well plate format (ISOLUTE® SLE+ 96-well plate, 820-0200-P01, Biotage), followed by separation on a Gemini, 50 mm × 2.0 mm I.D. 5 µm, column (00B-4435-B0, Phenomenex). Quantification was performed using selected reaction monitoring for the transitions m/z 739.5–>339.3 (DCV) and 747.5–>339.3 (isotope-labelled internal standard for DCV). The LLOQ was set to 1.64 ng/mL for DCV. A total of 9 quality control samples (3×low, 3×mid and 3×high concentration) were analysed within each batch of clinical samples (96-well plate), resulting in an accuracy of 2.46-2.62% RSE for DCV.

**PK/PD analysis**

Maximum concentration (C_max_) and time to reach maximum concentration (t_max_) was derived directly from the observed drug concentrations. The software’s automatic slope calculator was used to derive the elimination rate (λ) (adjusted R^2^ value with uniform weighting) and terminal elimination half-life (t_1/2_). The drug exposure measured as area under the concentration-time curve (AUC) was calculated for each drug using the trapezoidal method. Linear interpolation was used for acceding concentrations and log-linear interpolation for descending concentrations. For the individual analysis, the linear method was used for all measurements due to accumulation between the day 0 and day 28 measurements. Both the AUC to the last time point (AUC_last_, 8 hours for SOF and 24 hours for GS-331007 and DCV) and AUC to infinity (AUC_inf_) were calculated.

A non-compartmental pharmacodynamic analysis were conducted using viral load data from enrolment to day 14 to calculate area under the viral load – time curve and terminal elimination half-life of the viral clearance curve, using the same methodology as explained above. In addition, the relative reduction in viral load between enrolment and day 1, and between enrolment and day 7 were calculated. Ordinary linear regression of drug exposure (AUC_last_) from the individual pharmacokinetic analysis and the outcome measurements were performed in GraphPad Prism 9.3.1 (GraphPad Software, San Diego, California USA).

**Limitations of PK/PD analysis**

Two different sampling schedules were used to collect pharmacokinetic samples on day 0 and day 28, resulting in an overlapping sampling profile overall. Therefore, data collected within an individual on a specific day was not dense enough to justify a non-compartment analysis. However, if the data on day 0 and 28 were combined. it resulted in a complete pharmacokinetic profile for the individual. Two separate pharmacokinetic analyses were carried out. A naïve-pooled analysis was performed separately conducted on day 0 and 28 data. This resulted in a summary of exposure (AUC and Cmax) and half-life of the drugs, but it would not be possible to link these measurements to treatment outcome due to the different sampling strategies. Therefore, a second analysis was performed in which the data from day 0 and 28 were pooled for each individual. This resulted in complete pharmacokinetic profiles for each patient, and the pharmacokinetic-pharmacodynamic analysed demonstrated no significant relationship between drug exposure and treatment outcome. Drug accumulation was observed for the sofosbuvir metabolite GS-331007 and DCV between day 0 and 28. However, the individual analysis should still generate median exposure values for each patient, which can be linked with treatment outcome. Another drawback is that no pre-dose samples were collected. Therefore, the non-compartmental analysis will assume no concentration at day 0 on day 28 for GS-331007 and DCV, even though drug accumulation was observed.

**Appendix 1 figures**

**Appendix 1 - Figure 1: Mean (95% CI) HCV RNA by visit day**

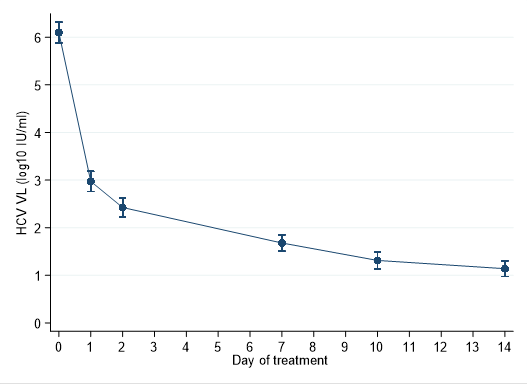

**Appendix 1 - Figure 2: time to viral suppression <LLOQ and eventual treatment outcome**

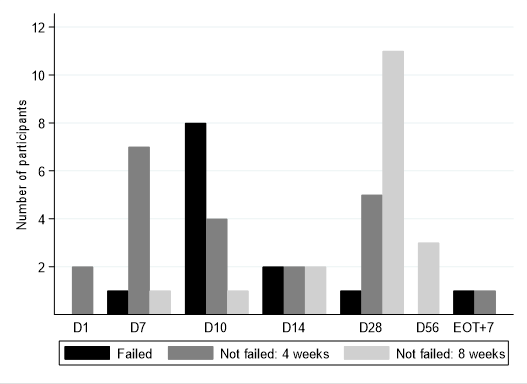

**No treatment failures in 8 week arm. D28 is the EOT visit for those who received 4 weeks. D56 visit is the EOT visit for those who received 8 weeks.*

**Appendix 1 - Figure 3: timing of treatment failure (confirmed HCV VL >2000 IU/mL) (n=13).**

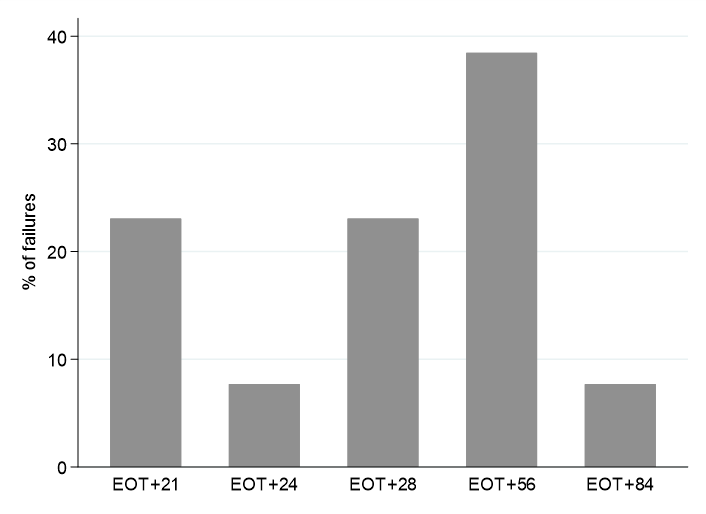

**Note twice weekly sampling in first 4 weeks after EOT, monthly thereafter.*

**Appendix 1 - Figure 4: All sofosbuvir-resistance associated substitutions at baseline (with treatment outcome)**

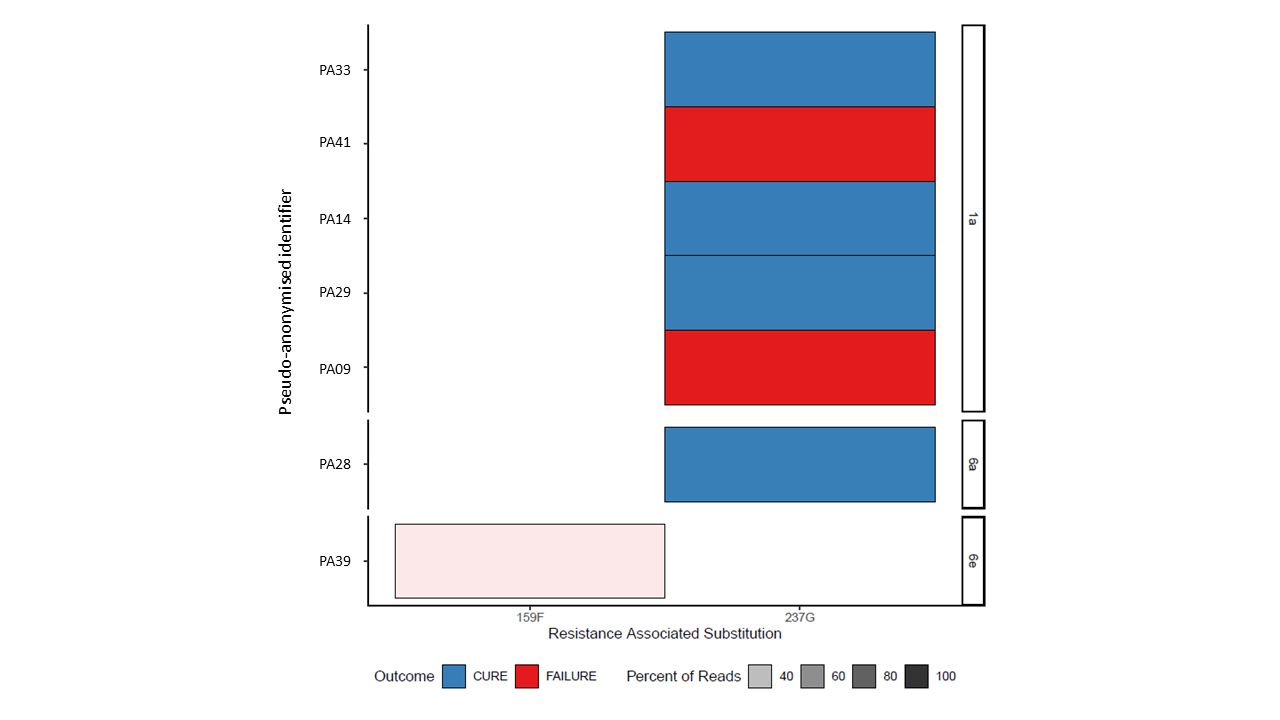

**Appendix 1 - Figure 5: Proportion of each subtype with sofosbuvir-resistance associated substitutions at baseline (with treatment outcome)**

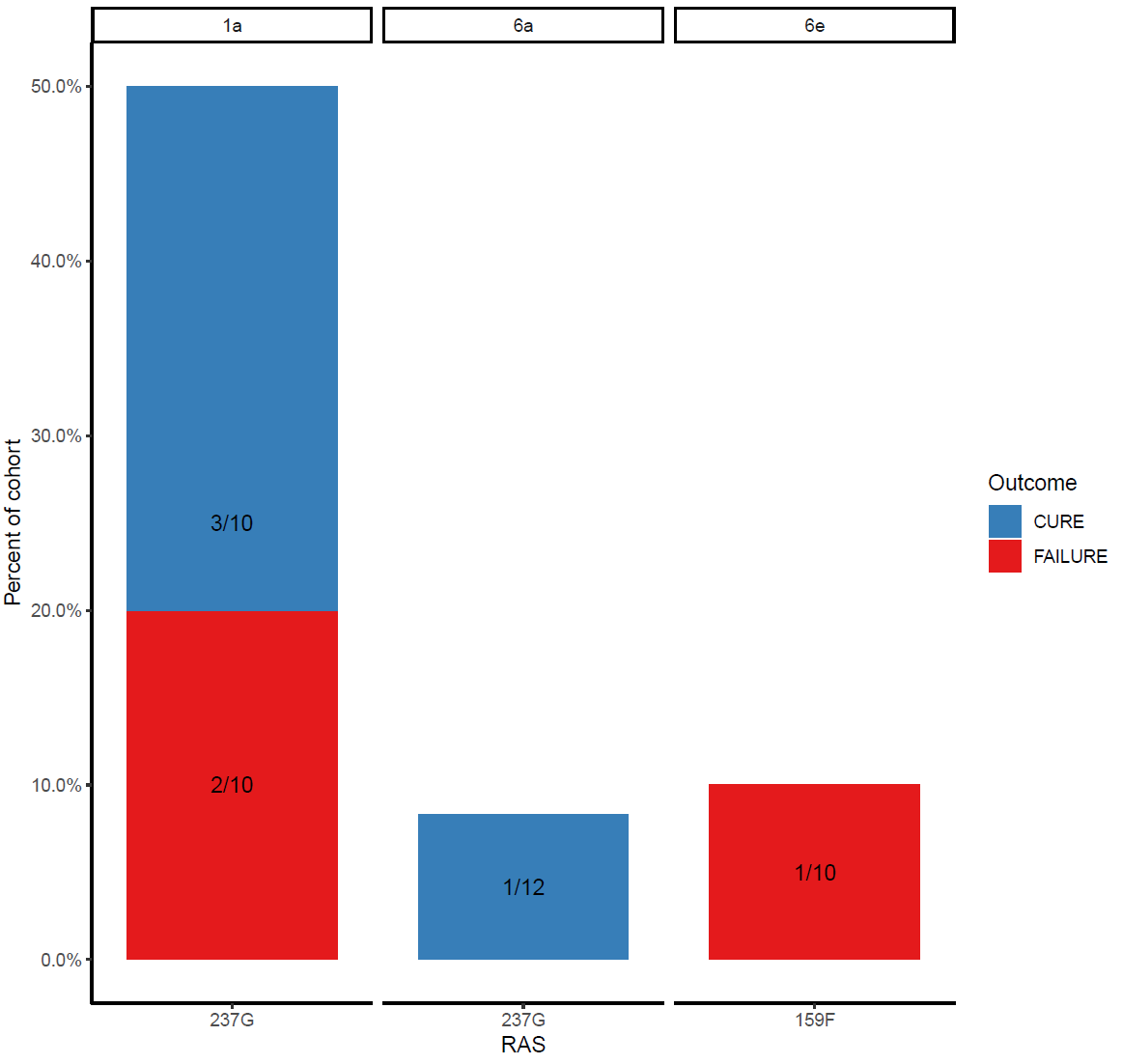

**Appendix 1 - Figure 6: All daclatasvir-resistance associated substitutions at baseline (with treatment outcome)**

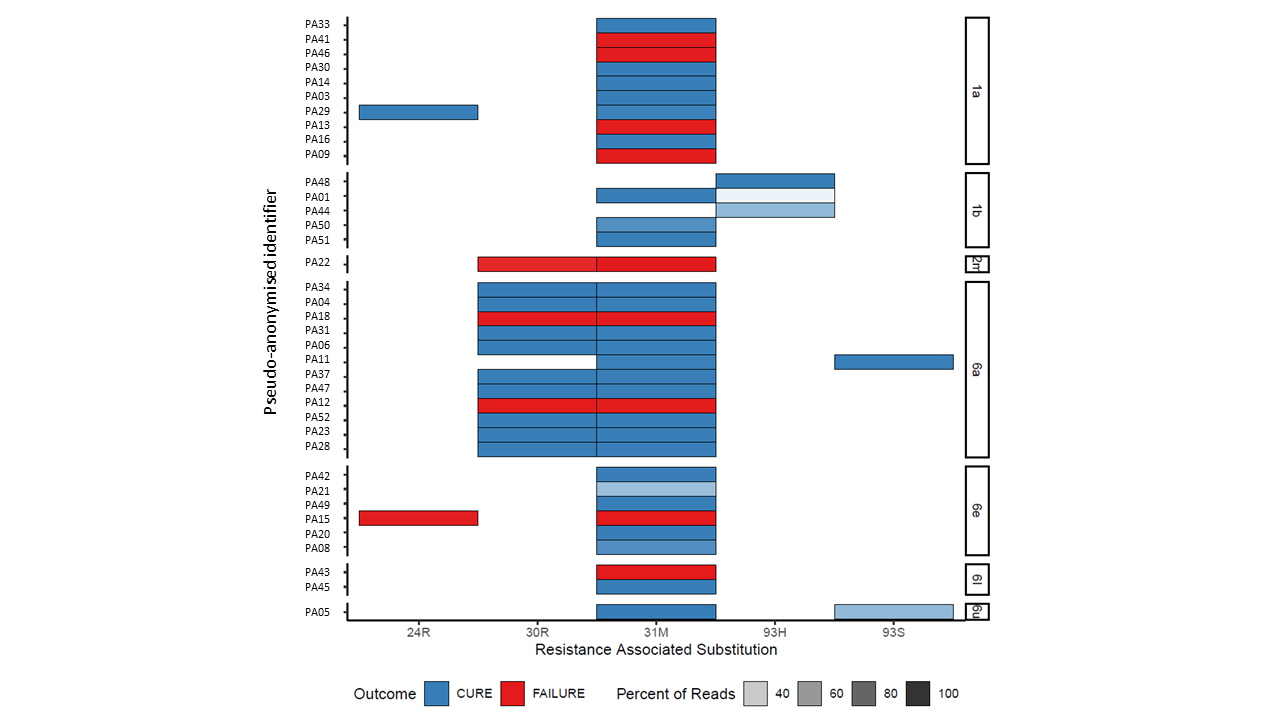

**Appendix 1 - Figure 7: Proportion of each subtype with daclatasvir-resistance associated substitutions at baseline (with treatment outcome)**

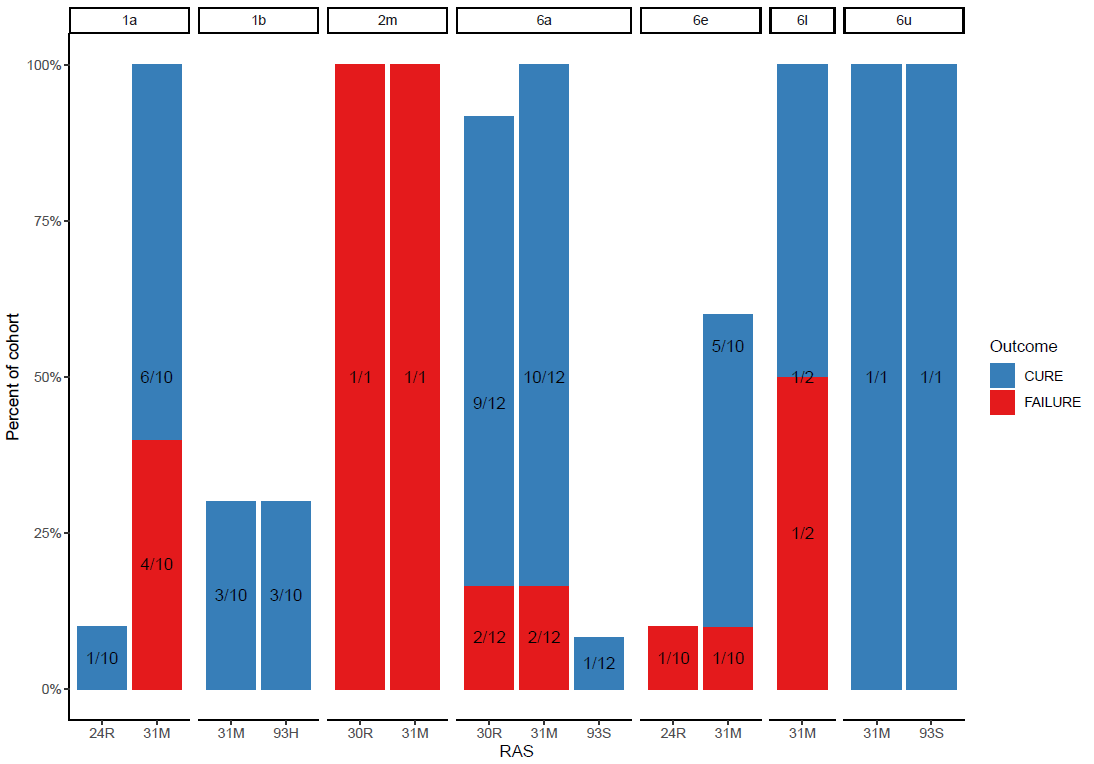

**Appendix 1 - Figure 8: SOF-RAS at baseline (D0), time of virological rebound (R) and start of retreatment (RT_W0)**

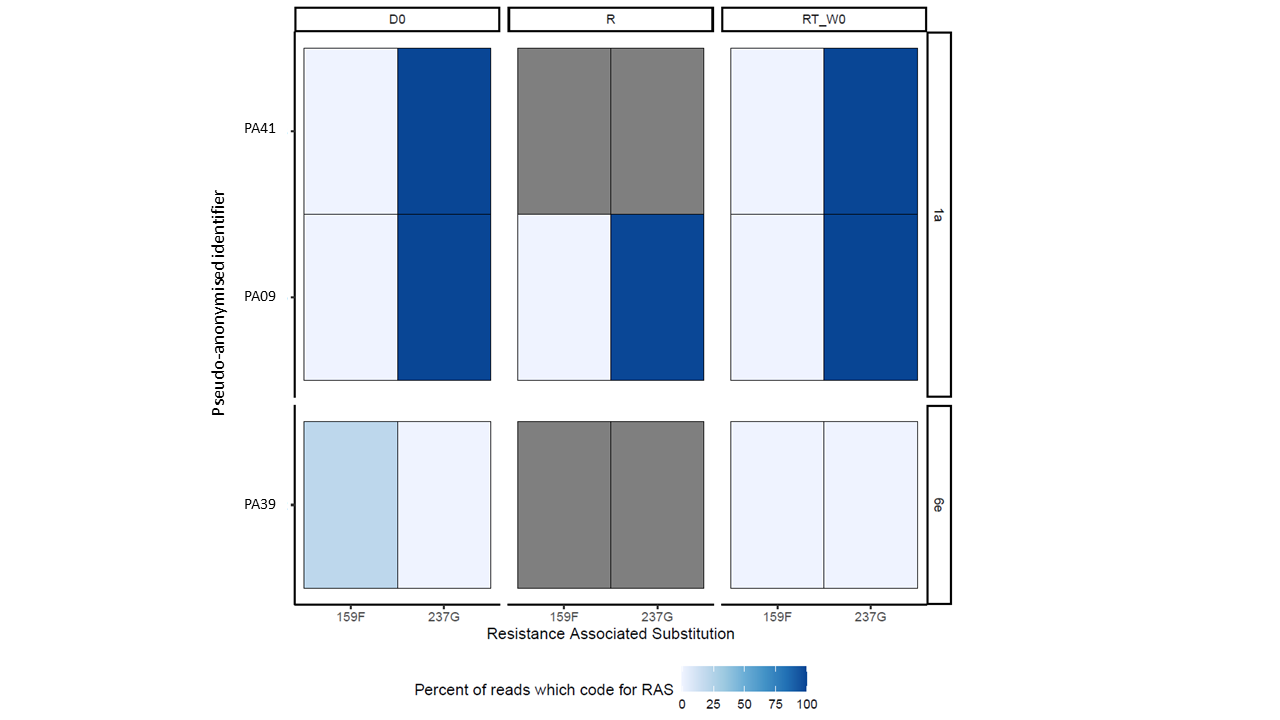

*Grey boxes represent missing data*

**Appendix 1 - Figure 9: DCV-RAS at baseline (D0), time of virological rebound (R) and start of retreatment (RT_W0)**

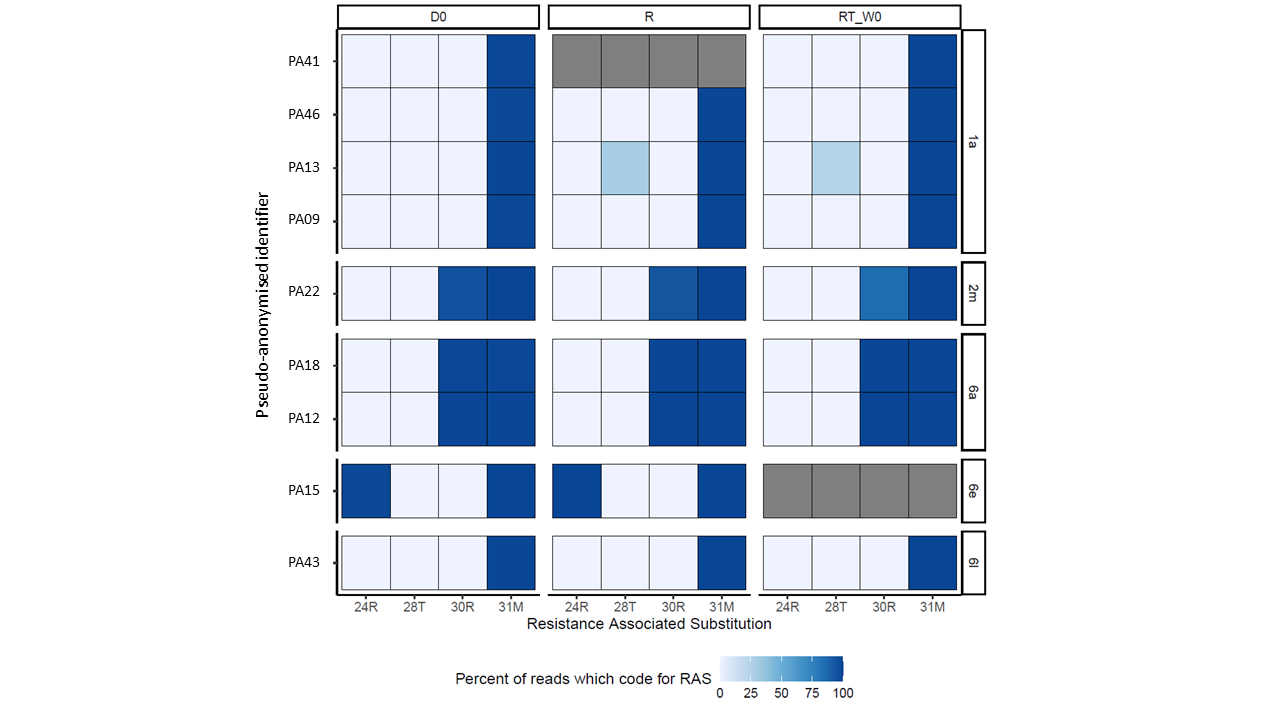

*Grey boxes represent missing data*

**Appendix 1 - table 1: Pharmacokinetic parameters from the naïve-pooled analysis**

|  | Sofosbuvir | | GS-331007 | | Daclatasvir | |
| --- | --- | --- | --- | --- | --- | --- |
|  | Day 0 | Day 28 | Day 0 | Day 28 | Day 0 | Day 28 |
| Cmax (ng/mL) | 1,320 | 1,070 | 988 | 1,230 | 1,170 | 1,110 |
| Tmax (h) | 1.00 | 1.00 | 3.00 | 4.00 | 3.00 | 3.00 |
| t1/2 (h) | 0.670 | 0.650 | 9.20 | 12.4 | 7.31 | 8.18 |
| AUClast (h×ng/mL)* | 1,550 | 1,600 | 10,500 | 14,600 | 11,400 | 12,400 |
| AUCINF (h×ng/mL)* | 1,550 | 1,600 | 12,700 | 20,400 | 12,800 | 14,400 |

*C_max_ is the maximum observed concentration, t_max_ is the time to reach the maximum concentration, t_1/2_ is the terminal elimination half-life (calculated using the 3-6 last concentration measurements, depending on drug and day), AUC_last_ is the total exposure to the last time point (8 hours for SOF and 24 hours for GS-331007 and DCV), AUC_inf_ is the total exposure extrapolated to infinity.*

**Extrapolation based on the last observed concentration measurement*

**Appendix 1 - table 2: Pharmacokinetic exposure from the individual analysis and pharmacodynamic parameters**

| Pharmacokinetics |  | |  |  |
| --- | --- | --- | --- | --- |
|  | Sofosbuvir | GS-331007 | | Daclatasvir |
| AUClast (h×ng/mL) | 1,140  (598-2,150) | 3,430  (2,200-4,720) | | 9,770  (5,080-16,200) |
| Pharmacodynamics |  |  | |  |
| AUC (days×IU/mL) | 252,000 (19,200-1,370,000) |  | |  |
| t1/2 (days) | 2.25 (0.986-5.22) |  | |  |
| %ReductionEnrolment-Day1 | 99.9 (99.0-100) |  | |  |
| %ReductionEnrolment-Day7 | 100 (100-100) |  | |  |

*Data is presented as median (5^th^ -95^th^ percentile). AUC_last_ is the total exposure to the last time point (8 hours for SOF and 24 hours for GS-331007 and DCV). AUC_14_ is the area under the viral load-time curve from enrolment (day 0) to day 14, t_1/2_ is the terminal viral half-life (estimated using at least three measurements), %Reduction_Enrolment-Day1_ is the reduction in viral load from enrolment to day 1, %Reduction_Enrolment-Day7_ is the reduction in viral load from enrolment to day 7.*

**The half-life could not be determined for one participant due to only one sample above the lower limit of quantification.*

**Appendix 1 - table 3: Pharmacokinetic-pharmacodynamic analysis**

| Linear regression |  |  |  |  |  |  |
| --- | --- | --- | --- | --- | --- | --- |
|  | Sofosbuvir |  | GS-331007 |  | Daclatasvir |  |
|  | Slope (95% CI) | *p* | Slope (95% CI) | *p* | Slope (95% CI) | p-value |
| Area under the viral load-time curve | -157  (-423 - 109) | 0.239 | 16.2  (-74.4 - 107) | 0.719 | -14.2  (-67.1 - 38.6) | 0.589 |
| Viral elimination half-life | 1.55×10^-4^  (-8.70×10^-4^ - 5.60×10^-4^) | 0.662 | -3.64×10-5  (-2.74×10^-4^ - 2.01×10^-4^) | 0.757 | 2.17×10^-5^  (-1.16×10^-4^ - 1.60×10^-4^) | 0.751 |
| Relative reduction in viral load at day 1 | 1.31×10^-6^  (-4.54×10^-6^ - 7.16×10^-6^) | 0.652 | 2.67×10-8  (-1.94×10^-6^ - 1.99×10^-6^) | 0.978 | 2.81×10^-7^  (-8.62×10^-7^ - 1.42×10^-6^) | 0.621 |
| Relative reduction in viral load at day 7 | 2.53×10^-7^  (-2.81×10^-7^ - 7.86×10^-7^) | 0.343 | 5.09×10-8  (-1.29×10^-7^ - 2.31×10^-7^) | 0.569 | 1.44*10^-8^  (-9.11×10^-8^ - 1.20×10^-7^) | 0.783 |

*95% CI is the 95% confidence interval around the slop*

**END OF DOCUMENT**
